# Predicting Dengue Incidence Leveraging Internet-Based Data Sources. A Case Study in 20 cities in Brazil

**DOI:** 10.1101/2020.10.21.20210948

**Authors:** Gal Koplewitz, Fred Lu, César Leonardo Clemente, Caroline Buckee, Mauricio Santillana

## Abstract

The dengue virus affects millions of people every year worldwide, causing large epidemic outbreaks that disrupt people’s lives and severely strain healthcare systems. In the absence of a reliable vaccine against it or an effective treatment to manage the illness in humans, most efforts to combat dengue infections have focused on preventing its vectors, mainly the Aedes aegypti mosquito, from flourishing across the world. These mosquito-control strategies need reliable disease activity surveillance systems to be deployed. Despite significant efforts to estimate dengue incidence using a variety of data sources and methods, little work has been done to understand the relative contribution of the different data sources to improved prediction. Additionally, scholarship on the topic had initially focused on prediction systems at the national- and state-levels, and much remains to be done at the finer spatial resolutions at which health policy interventions often occur. We develop a methodological framework to assess and compare dengue incidence estimates at the city level, and evaluate the performance of a collection of models on 20 different cities in Brazil. The data sources we use towards this end are weekly incidence counts from prior years (seasonal autoregressive terms), weekly-aggregated weather variables, and real-time internet search data. We find that both random forest-based models and LASSO regression-based models effectively leverage these multiple data sources to produce accurate predictions, and that while the performance between them is comparable on average, the former method produces fewer extreme outliers, and can thus be considered more robust. For real-time predictions that assume long delays (6-8 weeks) in the availability of epidemiological data, we find that real-time internet search data are the strongest predictors of dengue incidence, whereas for predictions that assume short delays (1-3 weeks), in which the error rate is halved (as measured by relative RMSE), short-term and seasonal autocorrelation are the dominant predictors. Despite the difficulties inherent to city-level prediction, our framework achieves meaningful and actionable estimates across cities with different demographic, geographic and epidemic characteristics.

**Author Summary:** As the incidence of infectious diseases like dengue continues to increase throughout the world, tracking their spread in real time poses a significant challenge to local and national health authorities. Accurate incidence data are often difficult to obtain as outbreaks emerge and unfold, both due the partial reach of serological surveillance (especially in rural areas), and due to delays in reporting, which result in post-hoc adjustments to what should have been real-time data. Thus, a range of ‘nowcasting’ tools have been developed to estimate disease trends, using different mathematical and statistical methodologies to fill the temporal data gap. Over the past several years, researchers have investigated how to best incorporate internet search data into predictive models, since these can be obtained in real-time. Still, most such models have been regression-based, and have tended to underperform in cases when epidemiological data are only available after long reporting delays. Moreover, in tropical countries, attention has increasingly turned from testing and applying models at the national level to models at higher spatial resolutions, such as states and cities. Here, we develop machine learning models based on both LASSO regression and on random forest ensembles, and proceed to apply and compare them across 20 cities in Brazil. We find that our methodology produces meaningful and actionable disease estimates at the city level with both underlying model classes, and that the two perform comparably across most metrics, although the ensemble method produces fewer outliers. We also compare model performance and the relative contribution of different data sources across diverse geographic, demographic and epidemic conditions.

## Introduction

The incidence of dengue has risen dramatically over the past few decades. With an estimated 100-400 million infections each year, dengue threatens roughly 3.9 billion people in 128 countries and poses a growing health and economic problem throughout the tropical and sub-tropical world.^1^ As climate change and urbanization intensify, the geographic range of dengue is expected to spread even further.^2^ Though the disease often manifests asymptomatically, severe cases can lead to hemorrhage, shock and death.^3^ In Brazil, which we examine in this paper, dengue has been endemic since 1986, and is today considered to be experiencing a “hyperendemic scenario,” in which both fatalities and severe cases are rising.^4, 5^ In the decades since 1986 over 40% of all dengue deaths in the country have been taken place in the Southeast region, but mortality from the disease has been reported in all but two of Brazil’s states.

Health services have strained to address the burden of dengue morbidity and mortality, in the regions where it is endemic, through a variety of means. Without a reliable vaccine or an effective treatment to manage the illness in humans, one effort, promoted by the World Health Organization (WHO), has aimed to achieve better early case detection. By focusing on improving epidemiological surveillance and attaining more timely identification of outbreaks, public health officials hope that preventive measures to reduce the spread of the disease can be used more effectively (vector control methods include, for example, the distribution of mosquito nets). However, effective real-time tracking of the spread of dengue – let alone prediction – has proven difficult. This is particularly evident in sprawling countries like Brazil, in which health resources are spread thin over a vast range of localities in which dengue is endemic. Governments typically rely on clinic-based reporting for case counts, but in Brazil (as in other countries) this information is often lagged in time and subject to post-hoc revisions, thus limiting the potential effectiveness of interventions.^6,7^ Thus, the development of data-informed tools for dengue surveillance which provide accurate case counts in real-time has increasingly become a priority.

The transmission dynamics of dengue and the time scales at which they occur lend themselves to tracking patterns of infection. In tropical environments, *Aedes aegypti* and *Ae. albopictus* mosquitoes can transmit dengue viruses within a week of infection. Once infected by a mosquito, a person can become ill within a week, and show symptoms for up to 10 days (other mosquitos can subsequently pick up dengue from an infected person within a 5-day window).^8, 9^ A range of external conditions have also been shown to affect dengue transmission. Among these are precipitation, temperature and other seasonal weather patterns, which influence the spread of the disease by affecting the development and lifespan of the dengue-carrying mosquitos.^10, 11, 12, 13, 14^ Additional factors include the human population density in a given town or region, as well as the degree to which various mosquito control efforts have been implemented by local health authorities.^15, 16^

Harnessing these various factors, a large number of models have been developed over the years in the attempt to forecast or nowcast dengue incidence (that is, to either predict future case counts or to accurately estimate current counts in real time). These range from compartmental mechanistic models, based on a set of differential equations, to statistical autoregressive models such as Seasonal Autoregressive Integrated Moving Average (SARIMA), which leverage both seasonal patterns and recent trends to produce disease estimates, to models based on various machine learning techniques.^17, 18, 19, 20, 21^ Over the past few years, search activity on internet search engines has increasingly been explored as a potential data source for these models. As internet access in the developing world increased, researchers have shown the potential of applying user activity data from search engines and social media to make predictive estimates of dengue incidence levels.^29, 34^

However, much of the work in this field has been done at the national or state levels, with models estimating disease incidence over vast geographical swaths with highly varying local conditions and rates of disease.^22^ At the city level, smaller population sizes and fewer reliable data sources makes modelling disease rates more technically challenging, as previous work at this resolution has shown.^23^ Still, while national- and state-level estimates are no doubt helpful, estimating incidence at the city-level can be uniquely useful to local and national health administrators (as well as to international health organizations) – for example, in guiding a more granular distribution of resources such as mosquito nets. In recent years, more attempts have been made to fill this gap and models for estimating disease incidence at the city level in a number of tropical countries have been developed.^21, 23^ In Brazil, a joint effort by academics and health officials has produced “InfoDengue,” a system for dengue surveillance at the city level which has been running since 2015.^24^ Using weather time-series data, case reports and information from social networks, InfoDengue produces a risk map and dengue incidence estimates.

Delays and inaccuracies in reported disease surveillance data are some of the key difficulties in detecting and monitoring epidemics, and a number of approaches, such as Bayesian hierarchical modelling and constrained P-spline smoothing, have been used by researchers in the attempt to account for these delays and the uncertainty they introduce.^25, 26, 27^ Other efforts to mitigate the effect of delays in reporting have sought to incorporate novel real-time data sources, such as Twitter activity, in order to improve nowcasting model performance.^22^ More recently, a comparative study has found that dengue incidence forecasts tended to do well in situational awareness late in the season, whereas early season forecasts needed improvement, and suggested the use of multiple-model ensemble approaches to improve accuracy, an approach that had previously shown promise.^28, 29^ When recently applied to data from Vietnam, this “superensemble” approach to probabilistic seasonal dengue forecasting was indeed shown to be more accurate, on average, than the models that comprised it.^30^ Another approach shown to improve forecasting performance in urban areas, in both mechanistic models and artificial neural networks, has been to incorporate human mobility data as features.^31^

### Our contribution

We seek to to estimate dengue activity at the city level up to 8 weeks ahead of the publication of epidemiological reports, and to identify the degree to which different sources of data contribute to the performance of these models. In examining cities with a range of demographic and geographic characteristics, as well as varying epidemic histories, we hope to point to the specific circumstances in which different data sources and the underlying models leveraging them perform best – and thus to suggest which model set-ups be used in practice in the future, in different epidemic scenarios. In order to achieve those goals, we extend methodological frameworks previously used for flu surveillance. We assess the predictive performance of a collection of models by comparing their estimates, produced in a strictly out-of-sample fashion (only using information that would have been available at the time of prediction), with the subsequently observed dengue incidence. The underlying statistical methods we compare are both regression-based (LASSO) and non-parametric ensembles (Random Forest), and the data sources we leverage for these estimates are: (a) weekly incidence counts from prior years (seasonal autoregressive terms), (b) weather measurements, and (c) real-time dengue-related Google Search Trends data. We evaluate the performance in tracking dengue in 20 cities in Brazil and highlight the conditions in which this framework achieves more accurate predictions. Our results show that despite the difficulties inherent to predictions at the city level, our framework achieves meaningful, actionable estimates, and highlights the conditions in which our models perform most accurately. Finally, we find that our approach is capable of identifying whether or not an upcoming season will experience an epidemic with accuracies above 75%, up to 8 weeks ahead of available reports.

## Materials and Methods

### Data

We used three distinct sources of information for our study: (a) historical dengue incidence from Brazil’s Ministry of Health, (b) Google search frequencies of dengue-related queries, aggregated at the state-level, for the states in which the 20 chosen cities are located, and (c) Weather data, obtained from the Modern-Era Retrospective analysis for Research and Applications, Version 2 (MERRA-2).^32^

We analyzed weekly dengue activity in 20 cities in Brazil: Aracaju, Barra Mansa, Barretos, Barueri, Belo Horizonte, Eunápolis, Guarujá, Ji Paraná, Juazeiro do Norte, Manaus, Maranguape, Parnaiba, Rio de Janeiro, Rondonópolis, Salvador, Santa Cruz do Capibaribe, São Gonçalo, São Luís, São Vicente, Sertãozinho, and Três Lagoas. We chose these Brazilian cities based on several criteria. First, they all had populations over 100,000 by July 2016 (the end of the time range we examined) and varied widely in population size above that threshold. Second, the cities were all chosen to be “dengue endemic” locations, experiencing between 7 and 10 epidemic years between 2001 and mid-2016 (following the definition of the Brazilian Ministry of Health, an epidemic year is one in which the number of confirmed cases of dengue fever exceeds 100 per 100,000 persons^33^). Finally, they were chosen from a wide geographic range of 13 different states in Brazil and have a wide range of population densities, both of which are epidemiological factors known to influence disease dynamics. For the full summary of the demographic and geographic characteristics of the different cities, see **Table B in the S1 text**.

### Epidemiological data

Weekly dengue case counts from January 2010 to July 2016 were obtained from the Ministry of Health of Brazil directly. We confirmed that the ministry-reported annual totals, which are based on a combination of PCR testing and syndromic diagnosis by local physicians and other health practitioners, match the sum of case counts over each year at the state level (as can be found on [the DataSUS service](http://www2.datasus.gov.br/DATASUS/index.php?area=0203&id=29878153)). Nevertheless, this observable data from reported cases likely underestimates the total number of cases, due to non-comprehensive testing, as well as cases that were diagnosed but ultimately not reported. This effect might vary through time and across different geographies.

### Online search volume data

Weekly Google search frequencies for dengue-related queries were obtained from Google Trends (www.google.com/trends) using the Google Health Trends API. The Google Trends API was accessed using the gtrends-tools interface (https://github.com/fl16180/gtrends-tools). The search terms were downloaded at the state-level, for the states in which each of the 20 cities is located (Google Trends data at the city-level are not currently available in Brazil).

For online search term selection, we initially sought to use Google Correlate (www.google.com/correlate), which is designed to identify search terms correlating highly with a given time series. This method has been used in the past with success.^34^ However, since most of the search terms returned by Google Correlate for our time series of dengue incidence were unrelated to dengue, and since it was discontinued in the course of our work (in December of 2019), we instead used the Google Trends (www.google.com/trends) tool to identify queries which are highly correlated with the term ‘dengue’ (a feature enabled by the Google Trends interface). In order to ensure the model was robust and generalizable, we ignored terms unrelated to dengue, and verified the terms with a native Portuguese speaker. The weekly aggregated search frequencies of these terms were then downloaded within the time period of interest. Importantly, since we intended the method to generalize to states and cities across Brazil, we used the same terms for the 20 cities. The query terms are presented in Table A in S1 text.

### Weather data

Weather data were collected from MERRA-2 (Modern Era Retrospective-analysis for Research and Applications). The MERRA-2 data are publicly available through the Global Modeling and Assimilation Office (GMAO) at NASA Goddard Space Flight Center. For each of the 20 cities, daily weather indicators from Jan 1 2000 to Dec 31 2016 were created, with the following features: mean daily 2-meter air temperature (K), precipitation (mm), mean daily wind speed (m/s), and 2-meter specific humidity (kg/kg, dimensionless). We calculated the total accumulated rainfall in a day (mm) as the sum of hourly precipitation (kg/m2/hr, which is equivalent to mm/hr) over the 24-hour period. These data were then aggregated into weekly reports, in the range of dates between January 2010 and July 2016, to align with the epidemiological dengue incidence data.

The weather data were produced at a naive resolution of 0.5 x 0.625 degrees, which works approximately to a ∼50 square km grid cell. Attributing these data to a specific city, then, involved overlaying the rectangular grid of weather data onto a spatial file outlining city boundaries, and taking the weighted average of grid cells covering the city boundary. Given the modelled nature of the MERRA-2 data, the data are never missing (there is full temporal coverage in the range of dates studied).

## Methods

Our model draws on a range of data sources that have been used in the multivariate linear regression modeling framework ARGO (AutoRegressive model with GOogle search queries as exogenous variables), previously used to track flu incidence using flu-related Google searches.^35^ But the underlying machine learning methodology in our model differs fundamentally, and we extend other aspects of previous models significantly. We introduce Random Forest-based prediction in addition to previously tested L1-based (LASSO) regularized regression models. This new model was used to combine information from historical dengue case counts and dengue-related Google search frequencies, as well as weather data, with the goal of estimating dengue activity at different time ranges ahead of the publication of official health reports.

At a high level, our models are re-trained each week on data available at the time of prediction in order to estimate an out-of-sample nowcast of dengue incidence for that week. The weekly generated training sets consisted of a growing time-window which contained incidence data from time points up to 8, 6, 3 or 1 weeks prior to the time of estimation. The minimal time-window used for a single point prediction contained 52 weekly data points (a full year), and the maximal time-window contained over 300, when estimating some of the final points in our range (in mid-2016). This growing window approach allowed the model to constantly improve its predictive ability by taking into account an ever-larger sample of the relationship between internet search behavior, weather, and dengue activity. An alternative approach, using a moving window of a constant size, proved to perform less well in most cases in our preliminary analyses. The initial target training data thus consisted of the 80 weekly case counts between January 1 2011 (the first point at which we had a full year of historical data) and June 30 2012, and this gradually expanding window of training data was used for point predictions 1, 3, 6 and 8 weeks in the future (see **figure 3** for an illustration of this). For completeness in our modeling approaches, we also incorporated information on dengue activity from one, two and three years before the time-to-prediction, to test if long-term seasonal activity would improve performance as the literature has suggested.^15, 29^

### Model formulation and assessment

Our models were based on the assumption that when there are more dengue cases, more dengue-related searches will be observed. This is formalized mathematically via a hidden Markov model, as explained in Yang et al, 2015.^29^

Assuming that epidemiological reports were available with different time delays ranging from 1 to 8 weeks, we constructed models that would only have access to the most recent information available at the time of prediction. Thus, our models incorporated historical information in the form of autoregressive features from the prior 52 weeks, if available, or from a reduced set depending on the assumed delay in the availability of epidemiological information. In other words, taking J to be the number of weeks for which we incorporate incidence data as autoregressive features, we defined four different set-ups: J_8_= {8, 9, …, 52}, J_6_= {6, 7, …, 52}, J_3_ = {3, 4, …, 52}, J_1_ = {1, 2, …, 52}. For J_8_the assumed delay in the receipt of epidemiological reports is 8 weeks, for J_6_the assumed delay is 6 weeks, and so on. These choices of J capture the influence of short-term fluctuations, which has been shown to be strongly predictive for dengue case counts.^29^

The effect of long-term seasonality is also considered, implicitly and explicitly, by the inclusion of our expanding training window strategy, which incorporates new training samples as more data is collected every week, and by explicitly including as predictors weeks 78, 104, and 156 whenever they were available (the case counts 1.5, 2 and 3 years before the point in time being estimated). Finally, we define K as the set of non-autoregressive features being used in a given model set-up, which includes Google Trends data and weather data.

## Model parameter estimation

### LASSO Regression

The Least Absolute Shrinkage and Selection Operator (LASSO) is a linear regression technique that minimizes the residual sum of squares subject to a L1 norm.

At a given time t, we estimate the log-transformed case counts *y*_*t*_, *y*_*t*_ = log (*c*_*t*_ + 1), to be

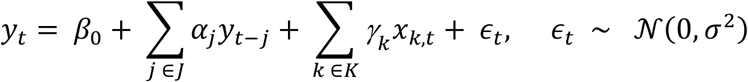

where *α*_*j*_ and *γ*_*k*_ are the estimation coefficients for *y*_*t − j*_, the observed dengue counts j weeks before the time t for which counts are being estimated, and *x*_*k,t*_, a given non-autoregressive feature *x*_*k*_ (such as a weather measurement or google trends search term) being used at time t in a given model set-up. *μ*_*y*_ is an intercept term and *∈*_*t*_ is the normally distributed error term. The L1 norm is a regularization technique that imposes a constraint over *α*_*j*_ and *γ*_*k*_, making the sum of the absolute value of the linear coefficients to not exceed a specific value (this value is a hyper-parameter, and is found via 5-fold cross validation). As a linear model, the coefficients associated with each feature are highly interpretable. L1 regularization also performs feature selection, zeroing out coefficients of features that contribute little to the predictions for each time window.

### Random Forests

Random Forests are a classification and regression method based on decision trees, models which can be used to approximate complex non-linear functions via simple partitions of the feature space. However, large and complex decision trees are prone to overfitting and high variance. This can be amended by using Random Forests, a form of bagging (“bootstrap aggregating”) in which multiple trees are trained on random samples of the training data – and then for a given input, the output is the averaged output of those trees.^36, 37^ To ensure the ensemble of decision trees is independent, for each split of each tree a random subset of predictors P’ is selected from the full set of predictors P. Finally, Random Forests have the advantage of being relatively interpretable, as widely accepted methods exist for calculating the relative importance of predictors in a “trained” forest (see ^[v]^, as well as ^38^). Still, they are not as intuitively interpretable as simple decision trees or linear models, in which one can more explicitly infer how the response variable changes in response to specific changes in features X.

All statistical analyses were performed with Python version 3.6.4 using Jupyter notebooks, using the statistical and machine learning libraries NumPy, Pandas, and Scikit-Learn. For both the LASSO regression and random forest-based models, the hypermeters (such as the alpha constant for LASSO or the maximum depth of the random forest) were set to the default values in the Scikit-Learn libraries, which were found to perform most consistently across our experiments.

#### Benchmark Models and Feature Sets

To our knowledge, few previous attempts were made to forecast or “nowcast” dengue incidence at the city level in Brazil. One such instance, which harnessed data from twitter to make estimates at both the country and city levels, found that tweets were useful for both forecasting and nowcasting dengue cases at the city level, though the association between the two was not as strong as at the country level.^25, 39^ Another such study focused on applying time-series analysis comparatively between two particular cities, Recife and Goiania, which have populations of a similar size.^40^ The Brazilian health authorities themselves typically release case counts 2-4 weeks after the fact, and frequently correct these figures substantially weeks after the initial publication. Thus, there was no clear external baseline with which to compare our results.

To evaluate performance with different assumptions about the availability of data and the relative contributions of various features, we constructed a number of internal benchmarks. First, we compared four different feature sets from our data sources: one solely with Google Trends data (which we label GT), a second solely with autoregressive data (AR), a third which included both (AR + GT), and a fourth that also took into account the weather data of each week and the week prior to it (AR + GT + W). In this way, we could assess the impact of each of the data sources at predictions with different models from different time horizons.

Second, we compared our two statistical methodologies, regression-based (LASSO) and non-parametric ensemble (Random Forest), and assessed how they performed relative to one another across the different feature sets and from different time horizons. In particular, we assessed the Random Forest model against the regression methodologies, which have been much better studied in the context of disease incidence nowcasting applications.

More generally, we evaluated which models and which data sources perform best at each time point with each methodology, while also summarizing performance across these in order to determine which methodology and feature set were most robust, and which led to the strongest performance across the board.

### Model assessment

We generated model estimates over the period between January 2011 and July 2016 with all of our models for each of the 20 cities, as selected following the previously described procedure. We used the following metrics to assess the performance of our models: root mean square error (RMSE), relative RMSE (R-RMSE), the R-squared coefficient of determination (R^2^) and the Pearson correlation coefficient. These were computed for the entire prediction period, over weekly intervals.

For each model, we also tested four variants based on simulating how recently the last official dengue case count report was received (denoted as 1, 3, 6, and 8-weeks before the “current,” predicted dengue report). Since the time delay between official case count reports is variable, it is important to assess how robust the models are to varying availability of autoregressive information.

Finally, to analyze more fully the long-term influence that historical dengue activity has on the future dynamics of outbreaks, we compared our selected AR model with an enhanced AR model, which included additional seasonal autoregressive features characterizing historical dengue activity (occurring up to 3 years in the past). Our results, which can be seen in **Figure A** and **Table C** of the **S1 text**, were effective in some cities but not in others, and so were not incorporated into the final model.

### Utilizing dengue activity point estimates to predict an incoming epidemic in Brazil

Building on the primary model for nowcasting real-time dengue incidence, we also tested our ability to predict, as a *binary* task, whether or not an epidemic would occur as a dengue season unfolds. More specifically, for each of the 20 cities, we assessed whether the cumulative number of dengue cases (that is, both the available reported epidemic observations and the disease estimates produced by our models) crossed a specified threshold value, referred to as the epidemic threshold, on a weekly basis. As the assumed delay in the availability of “observed” epidemiological information is up to 8 weeks, we substituted the 8 most recent weekly “missing” reports using our dengue point-estimates, and aggregated them along with the current “observed” available information as to increase our ability to predict a potential epidemic every week. Specifically, if the cumulative number of cases for a given time interval *t*_*e*_ exceeded the epidemic threshold value, we labelled the interval as epidemic. If it did not, we labelled it as non-epidemic. If our model using our substituted point estimates successfully predicted an epidemic within a dengue season as defined by the cumulative official case counts, we considered that season as a true positive. If the model did not predict an epidemic during all its weekly assessments and this remained consistent with the official epidemiological data, we considered that case a true negative. We generated the binary classification dataset by dividing the historical dengue activity time-series of each city into 52-week time intervals. These time intervals empirically center the high dengue activity periods, and keep the inter-outbreak activity (seasons with low dengue activity) at the start and the end of each interval. For each time interval, the cumulative dengue activity was calculated: from 0 in the first week, *t*_*0*_ to the total number of cases at week 52, or *t*_52_.

Given that the distribution of epidemic and non-epidemic intervals depends on the selection of the epidemic threshold – we tested and repeated our task using a range of values consistent with the standard thresholds reported in the literature, from 100/100,000 to 300/100,000.

## Results

When assuming short delays in the receipt of real dengue case count reports, we found that our models accurately estimated dengue incidence in 19 out of the 20 cities, across varying population sizes and local conditions. In the models in which the autoregressive case counts were included as features, a delay of one week in the receipt of real data resulted in an average error rate of under 0.5 relative RMSE. In this scenario, the model based only on Google Trends (GT) features underperforms relative to the ones in which autoregressive data were included, with performance around 0.85 in relative RMSE (see **Fig 1** and **Fig 2**).

**Figure 1.**
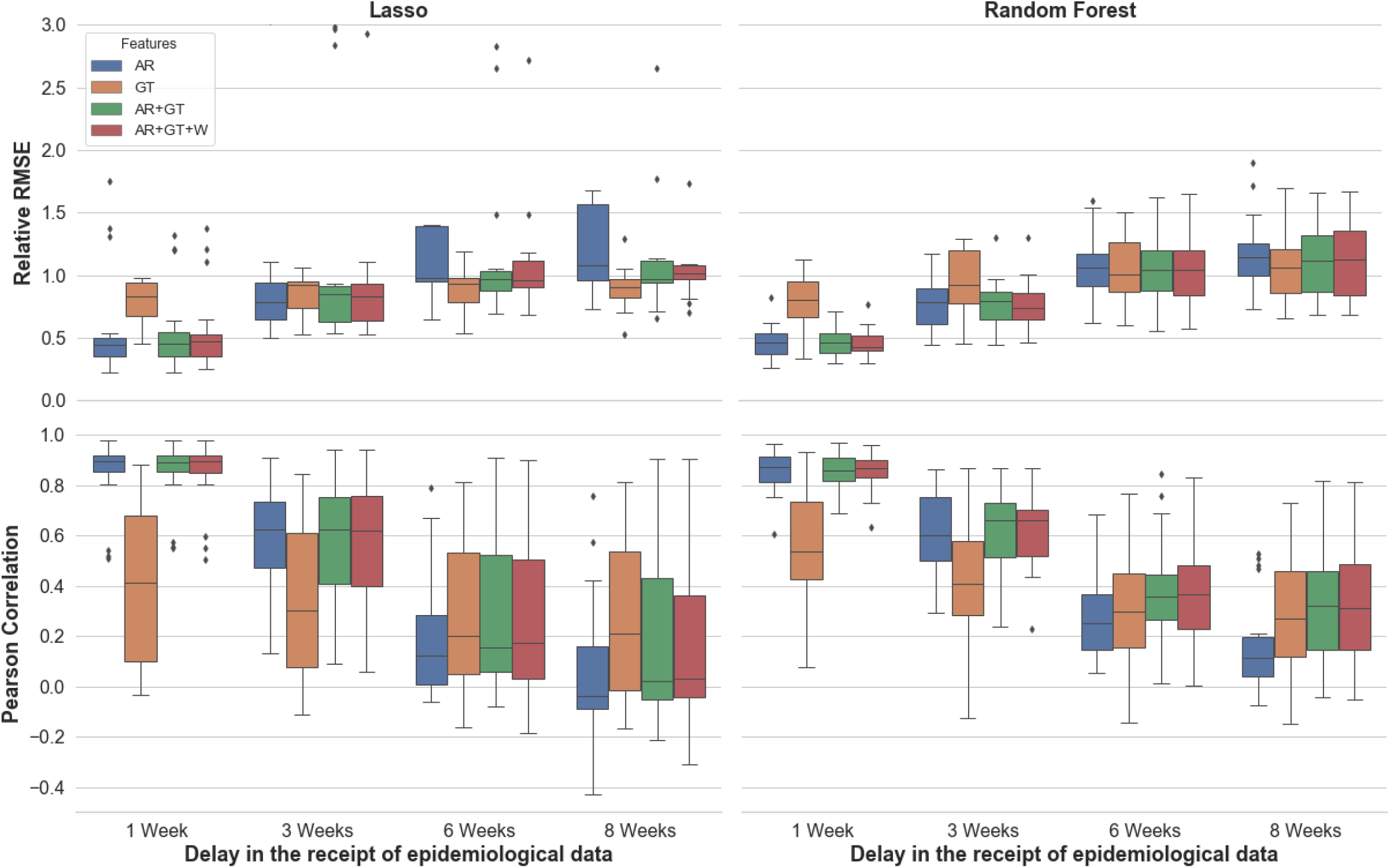
Performance across cities, as measured by Pearson Correlation and Relative RMSE. The colour of each box indicates the feature set used, and the x-axis notes the assumed delays in the receipt of epidemiological information. Each box shows the interquartile range of the metric for a given set-up (of feature set, assumed delay, and underlying model), while the whiskers show the rest of the distribution. Points beyond the whiskers in either direction are determined to be outliers

**Figure 2:**
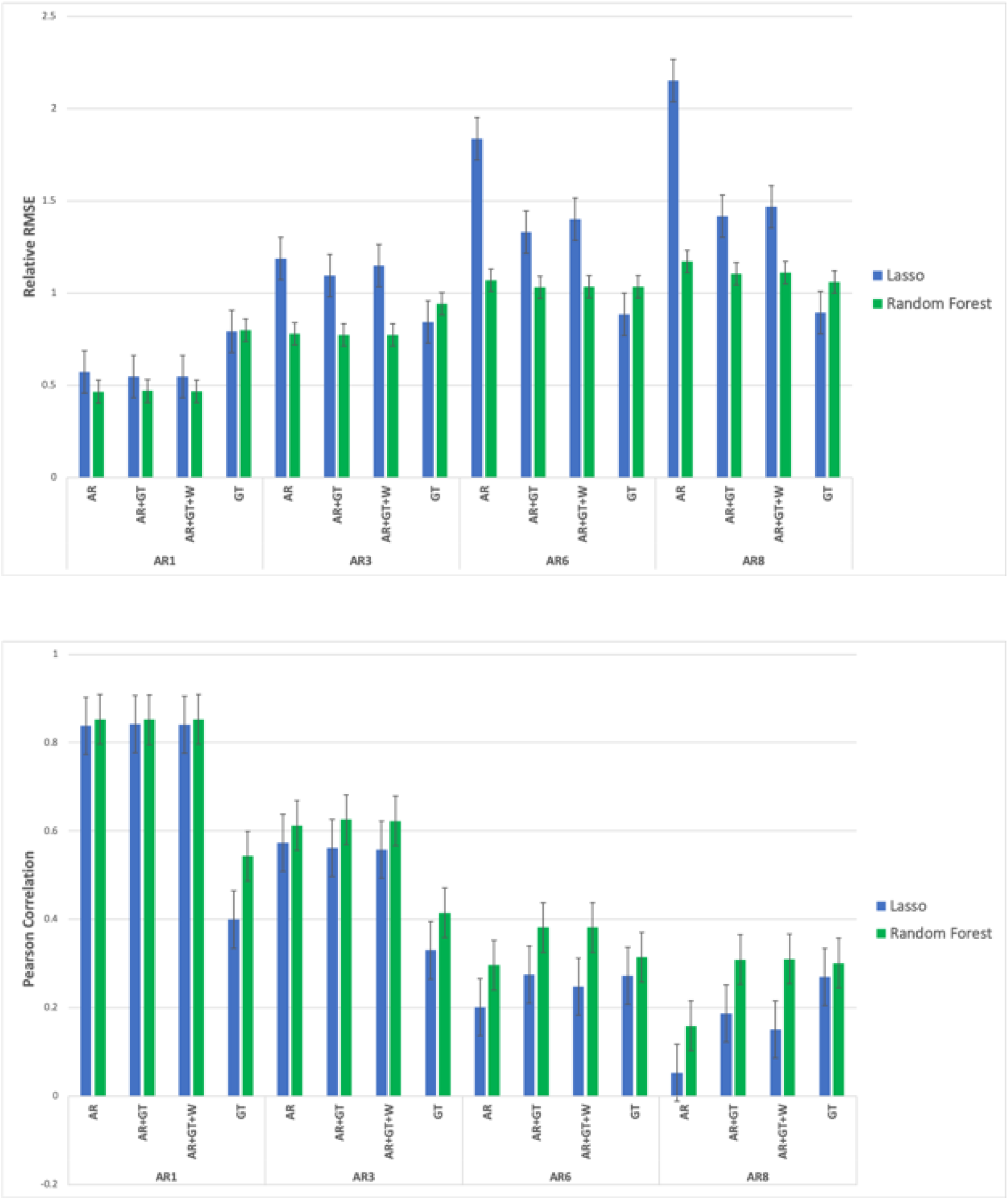
A comparison of Random Forest- and Lasso-based model performance. The mean is taken across the different cities, with the fill range of delays in availability of epidemiological information (from eight weeks, AR8, to one week, AR1) and the different feature sets (AR, GT AR+GT, AR+GT+W) shown.

When longer delays in the availability of epidemiological data are assumed, the LASSO-based model slightly outperformes the Random Forest-based models, and the best-performing feature set is GT. This advantage narrows in scenarios which assume shorter delays, of 1-3 weeks in advance, in which cases the two underlying methodologies tend to perform comparably. The Random Forest-Based model, however, is more robust to changes in features and assumptions about the availability of real-time epidemiological data. It also tends to produce fewer outlying, extreme values (see **Fig 1** and **Fig 2**).

As assumed delays in the availability of epidemiological data grow smaller, performance improves across the board, with lower RMSE and higher Pearson correlation observed in all models. For predictions that assume very short delays in the availability of epidemiological data, short-term and seasonal autocorrelation were key to improving estimates and captured a substantial amount of dengue variability. For predictions that assume longer delays, the real-time Google search trends data captured the most substantial amount of dengue variability. To highlight these effects, we examine a number of cities in the figures below, and focus on the model that tended to be most robust across different feature sets: the underlying RF methodology, with AR + GT feature set. In **Fig 3**, we show nowcasts in four cities using this model: Sao Luis, Belo Horizonte, Barra Mansa and Maranguape. These cities were chosen based on their different population sizes, peak epidemic rates, and weather patterns, and so demonstrate the comparative behaviour of the model across this range of demographic and geographic characteristics, as well as their epidemic histories (see **Table B** in S1 text for specific demographic and geographic statistics in each of the 20 cities).

**Figure 3.**
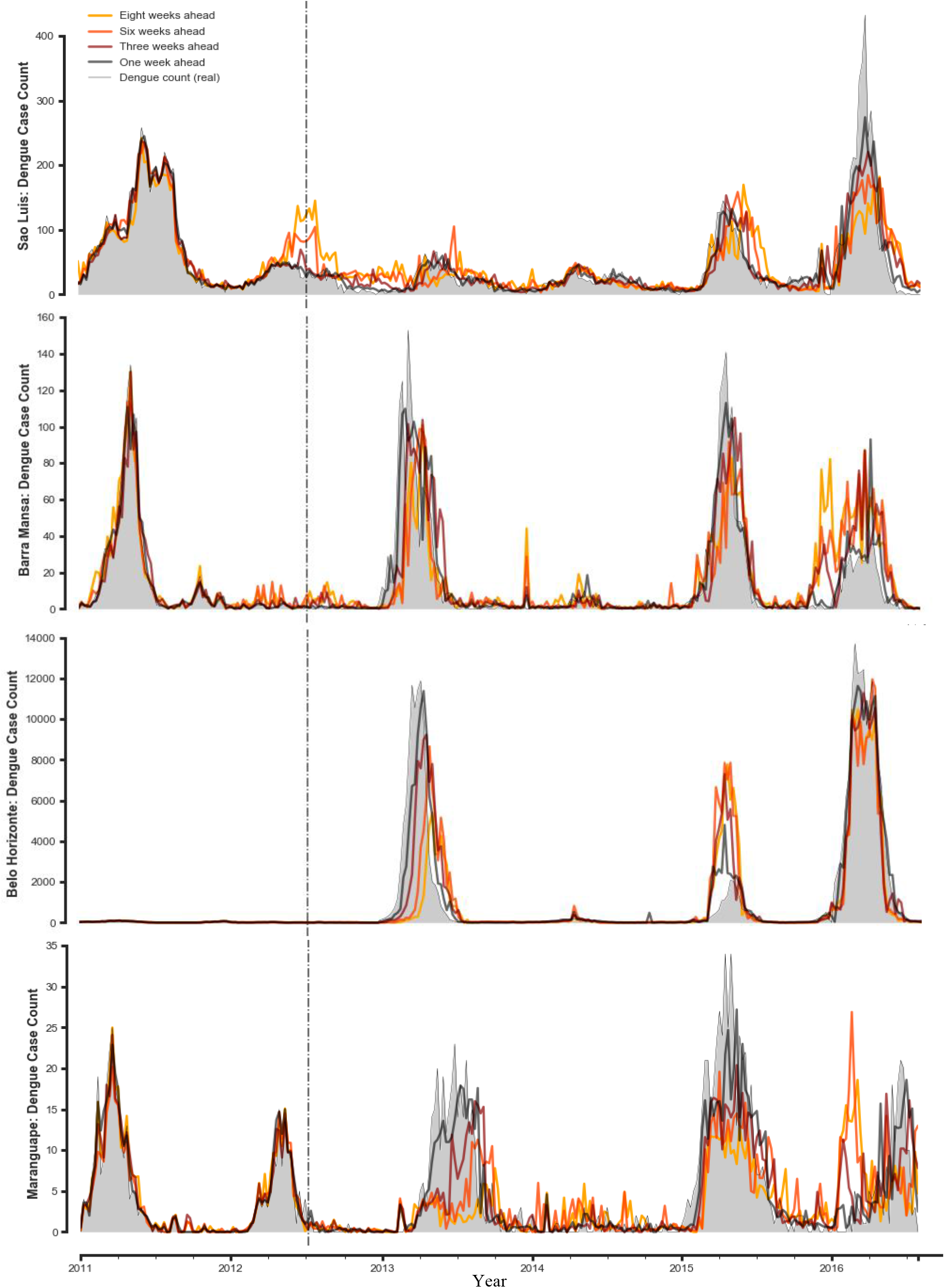
Dengue case estimates for 4 cities with different characteristics, as the delay in receipt of epidemiological data grows shorter.

To highlight performance at a more granular level and to allow comparisons between the different metrics, feature sets and the availability of epidemiological data, we focus on one of these, the city of Barra Mansa in the State of Rio de Janeiro. Barra Mansa was chosen because its density, area and population size are all close to the median of the 20 cities, and because its performance metrics and changes in the relative importance across model set-ups demonstrate some of the trends observed elsewhere (see **Table 1**). Data from all 20 cities are available at this resolution in **S2 spreadsheet**.

**Table 1.** Performance of dengue incidence prediction models from different time horizons, for the time period between January 2011 and July 2016, in the city of Barra Mansa, State of Rio de Janeiro, Brazil. Each time horizon is examined across all four possible features sets: autoregressive terms alone (AR), autoregressive terms together with Google Trends data (AR+GT) and with weather data (AR+GT+W), as well as google trends data alone (GT). Numbers in bold represent the best performance for a given model and autoregressive lag across each of the metrics. This corresponds to the **lowest** value for the **RMSE, relative RMSE** and **R^2** metrics, and the **highest** value for the **Pearson correlation** metric.

| Model | AR Lag | Features | RMSE | Relative RMSE | R<sup>2</sup> | Pearson Correlation |
| --- | --- | --- | --- | --- | --- | --- |
| Lasso Regression | 8 weeks | AR | 28.425 | 0.897 | 0.009 | 0.188 |
|  |  | GT | **23.606** | **0.745** | **0.317** | **0.563** |
|  |  | AR+GT | 23.828 | 0.752 | 0.304 | 0.555 |
|  |  | AR+GT+W | 24.67 | 0.778 | 0.254 | 0.505 |
|  | 6 weeks | AR | 26.65 | 0.845 | 0.122 | 0.355 |
|  |  | GT | 23.546 | 0.746 | 0.315 | 0.562 |
|  |  | AR+GT | **22.615** | **0.717** | **0.368** | **0.608** |
|  |  | AR+GT+W | 23.039 | 0.73 | 0.344 | 0.587 |
|  | 3 weeks | AR | 19.264 | 0.615 | 0.536 | 0.733 |
|  |  | GT | 21.581 | 0.689 | 0.418 | 0.649 |
|  |  | AR+GT | 18.859 | 0.602 | 0.556 | 0.752 |
|  |  | AR+GT+W | **18.789** | **0.6** | **0.559** | **0.753** |
|  | 1 week | AR | 12.485 | 0.4 | 0.804 | 0.897 |
|  |  | GT | 20.229 | 0.649 | 0.485 | 0.703 |
|  |  | AR+GT | 12.259 | 0.393 | 0.811 | **0.901** |
|  |  | AR+GT+W | **12.222** | **0.392** | **0.812** | **0.901** |
| Random Forest | 8 weeks | AR | 26.382 | 0.832 | 0.146 | 0.508 |
|  |  | GT | **21.061** | **0.664** | **0.456** | **0.68** |
|  |  | AR+GT | 23.355 | 0.737 | 0.331 | 0.596 |
|  |  | AR+GT+W | 22.514 | 0.71 | 0.378 | 0.631 |
|  | 6 weeks | AR | 24.625 | 0.781 | 0.251 | 0.591 |
|  |  | GT | 22.203 | 0.704 | 0.391 | 0.642 |
|  |  | AR+GT | 22.055 | 0.699 | 0.399 | 0.664 |
|  |  | AR+GT+W | **21.154** | **0.671** | **0.447** | **0.678** |
|  | 3 weeks | AR | 18.332 | 0.585 | 0.58 | 0.776 |
|  |  | GT | 21.07 | 0.673 | 0.445 | 0.676 |
|  |  | AR+GT | **17.613** | **0.562** | **0.612** | **0.793** |
|  |  | AR+GT+W | 19.354 | 0.618 | 0.532 | 0.749 |
|  | 1 week | AR | 11.047 | **0.354** | 0.846 | 0.92 |
|  |  | GT | 19.408 | 0.622 | 0.526 | 0.729 |
|  |  | AR+GT | **11.027** | **0.354** | **0.847** | **0.924** |
|  |  | AR+GT+W | 11.844 | 0.38 | 0.823 | 0.91 |

We also use the Barra Mansa to show the change in the relative importance of different predictors over time (see **Fig 4**). We observe that with an assumed delay of 8 weeks in the receipt of epidemiological data, Google search trends data tended to capture the greatest amount of variability, with some small amount also captured by some of the weather and autoregressive terms (**Fig 4**, left). With an assumed delay of 1 week in the availability of epidemiological data, however, the vast majority of the variability is captured by the first few autoregressive terms (**Fig 4**, right).

**Figure 4:**
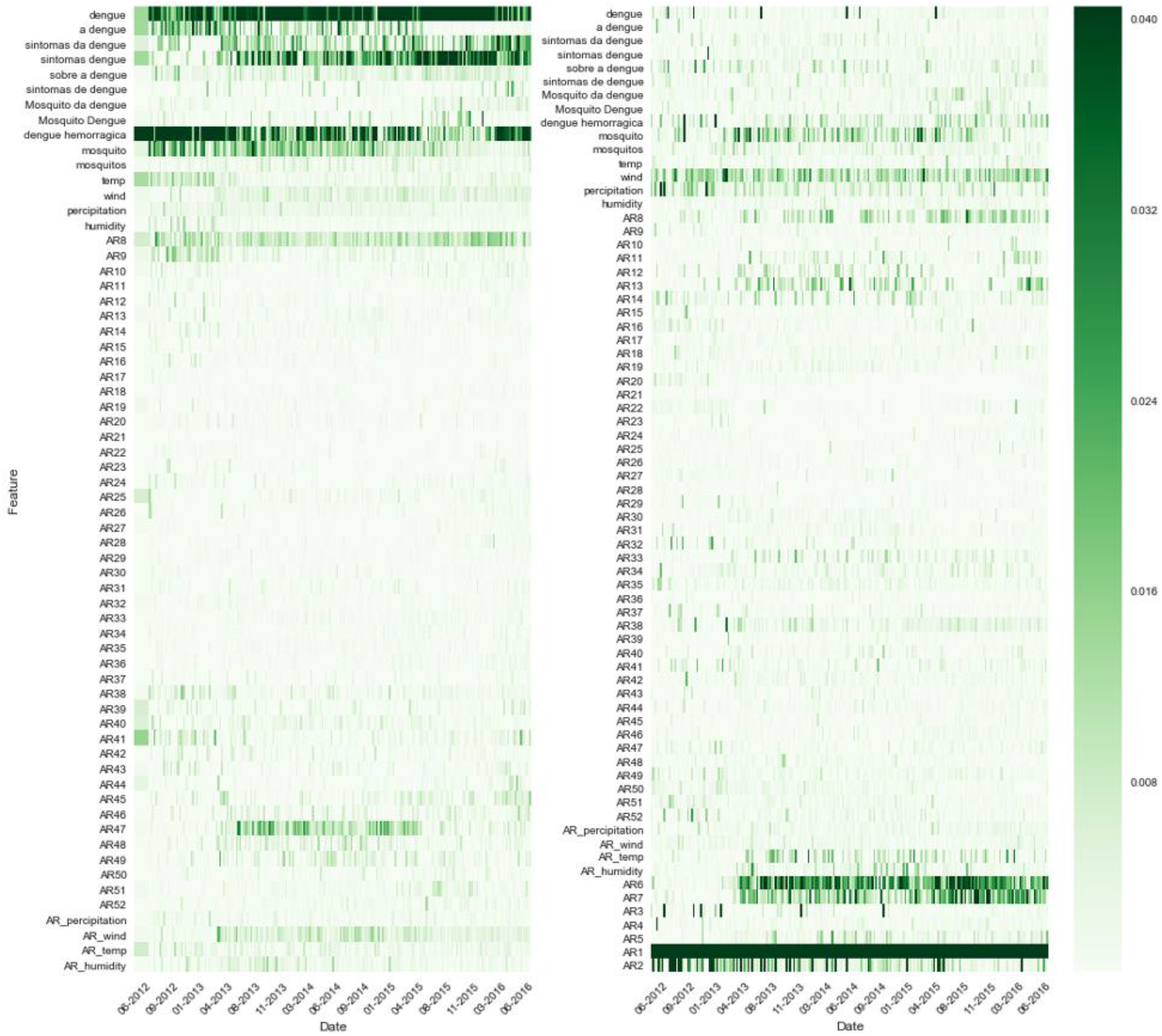
Change in the relative importance of different predictors over time. Barra Mansa, Random Forest model with full feature set (autoregressive epidemiological data, google trends data, and weather data). <u>Left:</u> An assumed delay of 8 weeks in the availability of epidemiological data. <u>Right:</u> An assumed delay of 1 week in the availability of epidemiological data.

In our analysis of the determinants of success of nowcasting at the city level, we find that long-term estimates tend to be more accurate when a city’s population is larger and when past dengue incidence has been relatively regular (see **Fig 5**, top right). We also plot success against the size and location of the city in Brazil (**Fig 5**, top left), and show that the decrease in prediction error, as the assumed delay in real-time information grows smaller, is consistent across the 20 cities (**Fig 5**, bottom).

**Figure 5:**
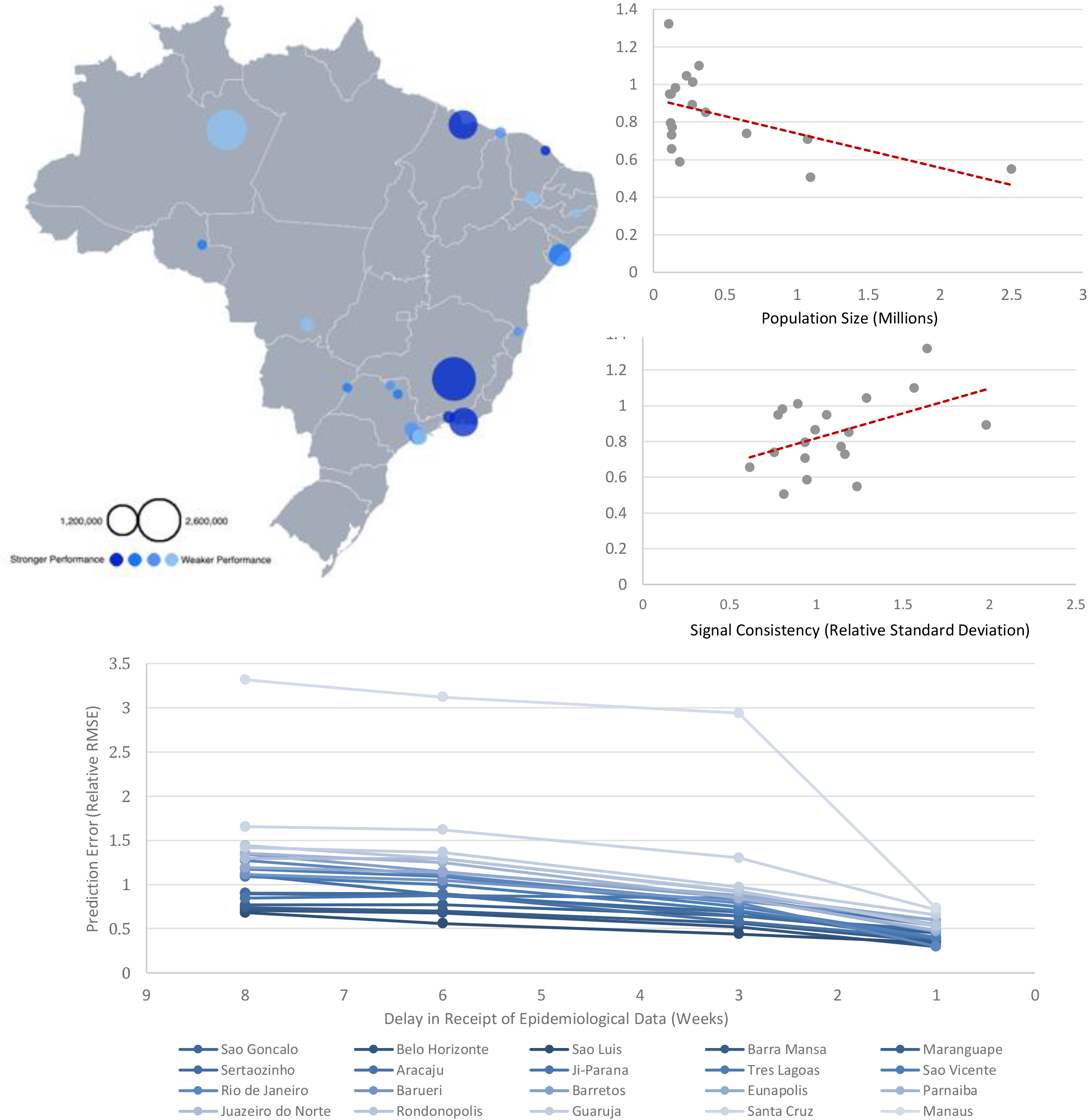
The determinants of success of nowcasting at the city level (random forest model, AR+GT feature set). **Top left**. City success, plotted on spatial map. The diameter of the circles reflects the size of the population, and a darker shade of blue indicates greater accuracy. **Top Right**. The effect of population size and dengue signal consistency on accuracy of predictions (averaged across the 20 cities). **Bottom**. Change in prediction accuracy (relative RMSE) as the delay in the receipt of epidemiological data grows shorter.

Finally, in the binary prediction task, in which we tried to predict in advance whether or not an epidemic would occur as a dengue season unfolds, we generated retrospective out-of-sample predictions using both the LASSO and the Random Forest methodologies, between October 5 of 2012 and July 31 of 2017, for the 20 cities in Brazil. The total number of time intervals generated were 60 (3 per city). To measure our model’s ability to predict an epidemic year, we utilized the standard definition of accuracy. We also measured the time difference Δ_*t*_ (in number of weeks) between *t*_*p*_, the week when our models nowcasted a dengue epidemic, and *t*_*e*_, the week in which the cumulative cases cross the epidemic threshold value. Δ_*t*_ is only measured for true positives (that is, in cases where *t*_*p*_ occurred earlier than *t*_*e*_). These metrics are summarized in **Figure B in the S1 Text. Figure C in the S1 Text** shows the distribution of epidemic and non-epidemic time intervals as a function of the epidemic threshold value. As the value of the epidemic threshold rises, the number of intervals classified as epidemic reduces, given the number of cumulative cases does not cross the threshold anymore.

Our results for the binary task show that our models are capable of successfully predicting epidemics, reaching accuracy values between .75 and .90, depending on the methodology and the type of information incorporated in the model. Lasso models achieve this with assumed delays in availability of “observed” epidemiological information of 5 to 7 weeks, whereas Random Forest-based models perform well with an assumed delay of up to 9 weeks. The choice of epidemic threshold does not affect these results.

## Discussion

Despite the difficulties inherent to predictions at finer spatial resolutions, our results show that our models and methodological framework for nowcasting dengue succeed at the city level and achieve accurate estimates. The conditions in which a given model set-up and chosen set of data sources perform best varies. While the LASSO-based model has a slight edge at predictions that assume a longer delay in the availability of epidemiological information, the random forest-based model produces fewer estimates with extremely high or low values, and can thus be considered more consistent and robust than the LASSO-based model (see and compare relative RMSE scores in **Fig 1** and in **Fig 2**). One possible reason for this is that tree-based models like random forests can capture non-linear relationships, which likely exist between at least a few of our features and dengue incidence counts. In the binary task, our outbreak detection addresses the concern that a simple majority-class predictor could achieve very high accuracy, by strongly outperforming the baseline, and see **Fig B in the S1 text** (in which the baseline is plotted as the grey line).

The predictive power of the different sources of information (epidemiological data, Google search data, and weather) used in this study varied depending on the expected delays of epidemiological data reports. For predictions that assume very short delays in the availability of epidemiological data, short-term and seasonal autocorrelation were key to improving estimates and captured a substantial amount of dengue variability while reducing the error rates, as measured by the R^2^ and relative RMSE metrics, respectively. For predictions that assume longer delays, the real-time Google search trends data captured the most substantial amount of dengue variability (see **Fig 4**). This is intuitively to be expected: the longer the span of time that has elapsed since observed data was available, the more useful the real-time proxy of Google Search Trends data becomes. Google Search Trends data also proved to be extremely effective in cases of sudden outbreaks, particularly when the scale was large enough. Such was the case with Barueri, a city in the state of São Paulo, in which there was a sharp spike in the number of dengue cases in 2015, well above peak incidence in previous years. In this instance, the feature set containing Google Search Trends data alone (GT) led to the most accurate performance at all time horizons, even when the assumed delay of epidemiological data was just a single week (see **S1 text, appendix**).

Weather data did not appear to have contributed significantly to the performance of the models (in the AR+GT+W set-up). This accords with previous work conducted on dengue case estimation, at the state level in Mexico, in which there was no significant uplift when temperature, relative humidity and precipitation were included in addition to the autoregressive terms.^15^ It seems, then, that for productionized autoregressive models deployed in real-time, the inclusion of weather data in addition to the case data and Google Trends data might not warrant the additional investment, if obtaining that data is in some circumstances is complex or expensive (this does not hold, of course, for models that are primarily dependent on climatic variables).

As noted above, we found that long-term estimates tend to be more accurate when a city’s population is larger and when past dengue incidence has been relatively regular (See **Fig 5, top right**). As Google Search Trends data can only be collected at the state level in Brazil, it is reasonable that its relevance to nowcasts made at the city level is higher in cases where the examined city’s population makes up a significant proportion of the state’s population, as in Rio de Janeiro, for example (or in cases where different cities in the state exhibit similar dengue incidence patterns). We also note that performance varies within a given city and model set-up, as the training window grows larger. As we can observe in figure 2, when predicting the first outbreak in a city’s epidemic, the estimates sometimes appear to lag the real counts by a week or so. Generally speaking, as the training window grows longer performance accuracy improves – but if outbreaks later in a city’s epidemic history are significantly weaker, the estimates sometimes overshoot, appearing to have “overlearned” the association between features and target from the previous outbreaks. In both the predictive and bi

Finally, though in some cities with certain characteristics the models perform better than in others, they tend to adapt quite well to the specific patterns of each city (lags, peak size of outbreak, etc.) after a period of training on a city’s past incidence data. Our framework contributes to the sparse but growing literature of infectious disease prediction models. Our results indicate that the lessons learned from dengue nowcasting in data-rich environments and at the country level can be generalized and tailored to track dengue in environments with significantly smaller populations, poorer data and a weaker disease signal. These insights can be leveraged towards future improvements in city-level nowcasting of infectious disease incidence.

On the whole, then, by accurately assessing suspected disease trends ahead of traditional disease surveillance systems – both in estimating case counts and in the binary task, in which performance significantly outstripped the baseline – this work can enable decision-makers to better plan for and implement dengue mitigation policies. These include scheduling education and mosquito control programs, informing supply chain efforts for medical supplies, and warning of outbreaks that are expected to be particularly severe. In particular, we hope the insights into the varying importance of features and the relative performance of model classes will be useful, as these vary in different circumstances – from the temporal offset at which real data is received by health professionals, to the variance in the geographic and demographic characteristics of the location being estimated.

### Further Work

One epidemiological feature to be included as input in future models is dengue incidence in proximate cities. Recent work has shown that certain geographical regions of Brazil have become increasingly vulnerable to dengue as transport infrastructure and other means of transportation to them has improved.^4^ Modelling this effect – for example, with cellular data, estimated volume of transportation, or simply with distance metrics – could improve estimates further, particularly for regions in which past observed case counts are less accurate or entirely unavailable. With the regression-based LASSO model, one naïve assumption that the relationship between the features and outcome variables is linear. This assumption is unlikely to be accurate (certainly across *all* variables), thus hampering model performance. But it could be that adding interaction and polynomial terms (which could then be narrowed down with a method like PCA) would improve LASSO performance, making it as robust as the Random Forest-based model, which does not assume linearity.

An additional promising direction is to design a composite model. This would take into account the finding that different feature sets, as well as the different underlying methodologies (LASSO and RF), led to the best performances in different cities and from different time lags. A composite model would incorporate these different sub-models and feature sets, and make use of them at the most fitting instances based on findings from the training data (for example, Google Search Trends data could be used as the feature set when making estimates that assume longer delays in the availability of observed case data). This could be constructed either explicitly based on rules or implicitly, and see the previously cited work on superensembles. To our knowledge, while superensembles have been used to estimate dengue incidence at the province level (in Vietnam), they have yet to be applied at the city level.^29, 30^

Finally, more refined hyperparameter tuning can lead to significant increases in performance for any one of the models and features sets set out above. There is a growing literature on efficient hyperparameter tuning techniques with ever-lower runtimes, and code libraries in which they are implemented could be easily deployed to increase the above models’ accuracy and reduce their error rates (including on custom metrics).

### Limitations

The weather data were produced at a naive resolution of 0.5 x 0.625 degrees, which works approximately to a ∼50 square km grid cell. Attributing these data to a specific city, then, involved overlaying the rectangular grid of weather data onto a spatial file outlining city boundaries, and taking the weighted average of grid cells covering the city boundary. Thus, there are some data fluctuations that come from grid cells that partially cover the ocean, or different altitudes/mountains. More generally, the approximations in data modelled and assimilated from MERRA tend to lead to less noise than weather station data (precisely because it is modelled) – so there are tighter but potentially less accurate oscillations in the time series.

Google Search Trends data are only currently available at the state level in Brazil. Were they to be made available at finer spatial resolutions, such as the city level (as they currently are in the United States) it is expected that performance would improve. This effect is likely to be particularly significant when making predictions that assume greater delays in the availability of epidemiological data, in which the Google Search Trends data were the most important features driving the forecast. Additionally, the process of selecting the Google search terms being tracked can be fine-tuned in the future, resulting in features that account for more of the variability in Dengue incidence.

It is likely that across the different cities we examined, different data collection methods are practiced, and that local public health officials have also introduced various health policy interventions. Both of these will have affected the consistency of the data across the 20 cities we examined, and will have introduced a degree of uncertainty. More generally, given that many cases are asymptomatic and that many symptomatic cases never get officially reported means that the “true” data are limited in scope to begin with. Thus, a central assumption of nowcasting studies such as this is that the reported, official dengue case counts (whether at the city level or at other resolutions) are at least a useful approximation of the underlying “true” incidence – and thus that estimating these reported counts is worthwhile.

Finally, it should be noted that the data we use have been subjected to “backfill.” That is, the dengue counts for a given week on which we trained our models are likely to have been subjected to post-hoc adjustments after they were initially reported in real-time. As such, this is a retrospective analysis, in which we use the finalized data, due to lack of availability of the original. In our experience, though, machine learning methods tend to learn patterns of missingness (for example, in flu forecasts), and so we expect it is likely that will be able to adapt to real-time predictions based on non-final data which has not been back-filled.^41^

## Supporting information

Results summary table

## Data Availability

All the data used for this study are either publicly available as reported in the manuscript, or available upon request.

## Acknowledgements

MS was partially supported by the National Institute of General Medical Sciences of the National Institutes of Health under Award Number R01GM130668. GK, CLC, CB, and MS thank the Harvard Data Science Initiative (US) for their support in the early stages of this project. The content is solely the responsibility of the authors and does not necessarily represent the official views of the National Institutes of Health. The authors thank Michael A. Johansson of the U.S. Centers for Disease Control and Prevention (CDC) for his insightful comments and contributions to this paper.

## Supporting Information

**S1 Text**. Supporting information text. This file includes: (1) Query terms used for Google Trends as Table A; (2) Demographic and geographic properties of chosen 20 Brazilian cities in Table B; (3) Dengue activity estimation using historical seasonality as Figure A, Figure B, Figure C, Table C and Table D. (4) Model performance across feature sets, cities and lags in the receipt of epidemiological information.

**S2 Text**. Measures of nowcasting performance across all models, features sets, and cities.

## Author Contributions

**Conceptualization:** GK MS CB.

**Data curation:** GK SM CLC.

**Formal analysis:** GK MS CLC.

**Funding acquisition:** MS.

**Investigation:** GK MS.

**Methodology:** GK FL CLC MS.

**Project administration:** MS.

**Software:**

**Supervision:** MS.

**Validation:** GK FL CLC MS.

**Visualization:** GK.

**Writing – original draft:** GK FL MS.

**All authors contributed and approved of the final version of the manuscript**.

