## Supplemental text 1 for "Predicting Dengue Incidence Leveraging Internet-Based Data Sources. A Case Study in 20 cities in Brazil": S1 Text Revised June 2021.docx

**Supporting Information (S1)**

Gal Koplewitz^1,2^, Fred Lu^3,4^, César Leonardo Clemente^2^, Caroline Buckee^2^, Mauricio Santillana^3,6, #^

^1^ Harvard J. A. Paulson School of Engineering and Applied Sciences, Cambridge, MA

^2^ Harvard T. H. Chan School of Public Health, Boston, MA

^3^ Computational Health Informatics Program, Boston Children’s Hospital, Boston, MA

^4^Department of Statistics, Stanford University

^6^ Department of Pediatrics, Harvard Medical School, Boston, MA

**Table A. Query terms used for cities, in Portuguese and English Translation**

| **Portuguese** | **English Translation** |
| --- | --- |
| dengue | dengue |
| a dengue | dengue |
| sintomas da dengue | dengue symptoms |
| sintomas dengue | symptoms dengue |
| sobre a dengue | about dengue |
| sintomas de dengue | dengue symptoms |
| mosquito da dengue | dengue mosquito |
| mosquito Dengue | mosquito dengue |
| dengue hemorragica | hemorrhagic dengue |
| mosquito | mosquito |
| mosquitos | mosquitos |

**Table B. Demographic and geographic properties of chosen 20 Brazilian cities.**

| **City Name** | **State** | **Population** | **Elevation** | **Density (2010)** | **Area (2017)** | | **Region** |
| --- | --- | --- | --- | --- | --- | --- | --- |
| Aracaju | Sergipe | 648939 | 4 | 3140.65 | 181.857 | Nordeste | |
| Barra Mansa | Rio de Janeiro | 183976 | 389 | 324.94 | 547.133 | Sudeste | |
| Barretos | São Paulo | 121344 | 530 | 71.6 | 1566.161 | Sudeste | |
| Barueri | São Paulo | 271306 | 719 | 3665.21 | 65.701 | Sudeste | |
| Belo Horizonte | Minas Gerais | 2501576 | 760 | 7167 | 331.401 | Sudeste | |
| Eunápolis | Bahia | 112318 | 189 | 84.97 | 1425.968 | Nordeste | |
| Guarujá | São Paulo | 318107 | 4 | 2026.8 | 144.794 | Sudeste | |
| Ji.Paraná | Rondônia | 127907 | 170 | 16.91 | 6896.649 | Norte | |
| Juazeiro do Norte | Ceará | 271926 | 350 | 1004.45 | 248.832 | Nordeste | |
| Manaus | Amazonas | 2145444 | 92 | 158.06 | 11401.092 | Norte | |
| Maranguape | Ceará | 127098 | 68 | 192.19 | 590.873 | Nordeste | |
| Parnaíba | Piauí | 152653 | 5 | 334.51 | 434.229 | Nordeste | |
| Rio de Janeiro | Rio de Janeiro | 6688927 | 2 | 5265.82 | 1200.177 | Sudeste | |
| Rondonópolis | Mato Grosso | 228857 | 227 | 47 | 4686.622 | Centro-Oeste | |
| Santa Cruz do Capibaribe | Pernambuco | 105936 | 438 | 261.2 | 335.309 | Nordeste | |
| São Gonçalo | Rio de Janeiro | 1077687 | 19 | 4035.9 | 248.319 | Sudeste | |
| São Luís | Maranhão | 1094667 | 4 | 1215.69 | 834.827 | Nordeste | |
| São Vicente | São Paulo | 363173 | 6 | 2247.88 | 148.1 | Sudeste | |
| Sertãozinho | São Paulo | 124453 | 579 | 273.22 | 403.089 | Sudeste | |
| Três Lagoas | Mato Grosso do Sul | 119465 | 319 | 9.97 | 10206.949 | Centro-Oeste | |

**Figure A. Improving dengue activity estimation using historical seasonality in Brazil municipalities**

To analyze the long-term influence that historical dengue activity has on the future dynamics of outbreaks, we conducted a study to compare our selected AR model and an enhanced AR model including key seasonal autoregressive features that characterize the historical dengue activity occurred up to 3 years in the past. We produced retrospective out-of-sample dengue activity estimates from January 4 of 2014, to July 06 of 2016, and used the RMSE metric as a benchmark against which to compare the performance of each model.

Our results, as presented in **Table C** below, show that the improvement that stems from the incorporation of seasonal trends in our autoregressive model was not present in every city and, in some cases, hindered the model’s predictive power. Given the unexpected results, we further investigated as to why the incorporation of seasonality was not effective in some cities, by conducting an autocorrelation analysis where the most recent historical dengue activity for each city was correlated with its own trend – from 1 week, up to 3 years in time-shift. We also opted to vary the size of the dengue incidence trend (from a vector of 52 weeks up to 156 weeks) to be correlated, with the purpose of obtaining autocorrelation values that are consistent with the expanding time-window used for our training methodology. The results of our autocorrelation, visualized in **Figure A** below, suggest that the length of the training periods used greatly affected the autocorrelation values of each AR lag, and could play a major role in the understanding and effective incorporation of seasonal trends used in our models.

**Figure B** shows the distribution of epidemic and non-epidemic time intervals as a function of the epidemic threshold value. As the value of the epidemic threshold rises, the number of intervals classified as epidemic reduces, given the number of cumulative cases does not cross the threshold anymore.

Unfortunately, the size of our dataset posed a big limitation in our seasonality study. We believe that as more epidemiological data becomes available, more extensive analysis could be conducted to further understand the relative predictive power of seasonality (specifically, in the case of municipalities in Brazil).

**Table C. Comparative effect of seasonality (AR vs. S+AR) as measured by RMSE, for all 20 cities over the different assumed delays in the receipt of epidemiological information. Models with lower RMSE are highlighted in bold for each time horizon.**

| **Delay in Receipt of Epidemiological Information** | **City** | **AR** | **S+AR** | **City** | **AR** | **S+AR** |
| --- | --- | --- | --- | --- | --- | --- |
| 1 week | Aracaju | **13.12** | 14.204 | Maranguape | **3.364** | 3.601 |
| 3 week |  | **19.953** | 22.788 |  | **4.374** | 5.032 |
| 6 week |  | **26.801** | 30.143 |  | **5.884** | 6.375 |
| 8 week |  | **28.555** | 30.943 |  | **6.853** | 8.388 |
| 1 week | Barra Mansa | **9.838** | 10.11 | Parnaiba | **5.266** | 5.283 |
| 3 week |  | **17.574** | 18.199 |  | **5.875** | 5.947 |
| 6 week |  | **24.117** | 26.533 |  | 7.236 | **6.133** |
| 8 week |  | **25.812** | 26.464 |  | 7.602 | **6.363** |
| 1 week | Barretos | **15.94** | 19.207 | Rio de Janeiro | 193.226 | **188.286** |
| 3 week |  | **27.029** | 39.1 |  | **490.189** | 578.742 |
| 6 week |  | **31.504** | 68.993 |  | 951.166 | **591.084** |
| 8 week |  | **36.964** | 90.771 |  | 1148.836 | **592.783** |
| 1 week | Barueri | 290.344 | **157.628** | Rondonopolis | **4.532** | 8.309 |
| 3 week |  | 740.518 | **365.142** |  | **6.481** | 15.733 |
| 6 week |  | 949.557 | **771.659** |  | **9.736** | 18.774 |
| 8 week |  | 1024.359 | **913.986** |  | **11.192** | 20.311 |
| 1 week | Belo Horizonte | **655.273** | 687.127 | Santa Cruz do Capibaribe | 14.979 | **7.006** |
| 3 week |  | **1395.979** | 2040.588 |  | 9.88 | **9.743** |
| 6 week |  | 2506.477 | **2470.836** |  | 10.306 | **10.139** |
| 8 week |  | 3226.541 | **2201.558** |  | **10.519** | 10.56 |
| 1 week | Eunapolis | **3.848** | 3.89 | Sao Goncalo | **81.258** | 120.44 |
| 3 week |  | **5.97** | 6.097 |  | **210.696** | 353.553 |
| 6 week |  | 8.455 | **7.409** |  | **472.3** | 745.077 |
| 8 week |  | 8.542 | **7.318** |  | **495.589** | 890.717 |
| 1 week | Guaruja | **12.375** | 13.412 | Sao Luis | **28.261** | 32.666 |
| 3 week |  | 20.769 | **19.774** |  | **47.538** | 55.685 |
| 6 week |  | 27.935 | **27.089** |  | **63.017** | 68.614 |
| 8 week |  | 30.781 | **30.473** |  | **67.744** | 73.94 |
| 1 week | Ji Parana | **10.053** | 10.411 | Sao Vicente | **20.441** | 33.968 |
| 3 week |  | 23.696 | **20.833** |  | **34.464** | 49.17 |
| 6 week |  | 23.161 | **22.653** |  | **49.027** | 80.82 |
| 8 week |  | 23.216 | **22.779** |  | **53.337** | 89.164 |
| 1 week | Juazeiro do Norte | **15.674** | 20.112 | Sertaozinho | **40.205** | 58.381 |
| 3 week |  | **24.602** | 32.972 |  | 65.878 | **49.943** |
| 6 week |  | **29.108** | 30.035 |  | **58.684** | 74.551 |
| 8 week |  | **29.542** | 29.937 |  | **70.249** | 76.268 |
| 1 week | Manaus | 39.382 | **35.95** | Tres Lagoas | **12.874** | 15.241 |
| 3 week |  | 111.976 | **67.639** |  | **22.404** | 45.357 |
| 6 week |  | 246.807 | **103.177** |  | **33.732** | 89.371 |
| 8 week |  | 305.165 | **122.868** |  | **30.527** | 81.872 |

**Table D:** Relative RMSE, Relative Bias, Relative Sharpness and Pearson correlation, over all 20 cities, across all features sets.

| **Feature Set\Metric** | **Relative RMSE** | **Relative Bias** | **Relative Sharpness** | **Pearson Correlation** |
| --- | --- | --- | --- | --- |
| **AR1** | 0.541 | -0.039 | 0.538 | 0.85 |
| **AR3** | 1.121 | -0.113 | 1.109 | 0.601 |
| **AR6** | 1.709 | -0.216 | 1.682 | 0.271 |
| **AR8** | 1.988 | -0.27 | 1.953 | 0.167 |
| **AR1 + GT** | 0.51 | -0.039 | 0.508 | 0.857 |
| **AR3 + GT** | 1.008 | -0.102 | 0.996 | 0.603 |
| **AR6 + GT** | 1.2 | -0.123 | 1.18 | 0.384 |
| **AR8 + GT** | 1.268 | -0.123 | 1.247 | 0.323 |
| **AR1 + GT + W** | 0.509 | -0.038 | 0.506 | 0.856 |
| **AR3 + GT + W** | 1.057 | -0.108 | 1.046 | 0.6 |
| **AR6 + GT + W** | 1.264 | -0.129 | 1.245 | 0.369 |
| **AR8 + GT + W** | 1.317 | -0.128 | 1.297 | 0.299 |
| **GT1** | 0.747 | -0.009 | 0.741 | 0.478 |
| **GT3** | 0.788 | -0.009 | 0.781 | 0.421 |
| **GT6** | 0.818 | -0.008 | 0.809 | 0.371 |
| **GT8** | 0.821 | -0.009 | 0.812 | 0.361 |

**Figure A. Autocorrelation plots for each of the 20 cities. Ordered from left to right, each column represents an autocorrelation plot for the city using:**

**a) A 52-week long autocorrelation**

**b) A 104-week long autocorrelation**

**c) A 156-week long autocorrelation**

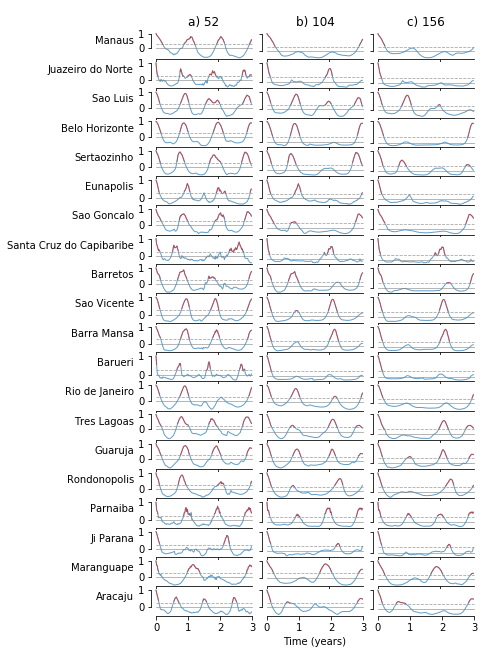

**Figure B. Line plots showing the accuracy and time difference** $\boldsymbol{\Delta}_{\boldsymbol{t}}$ **(in number of weeks) between** $\boldsymbol{t}_{\boldsymbol{p}}$**, the week when our models nowcasted a dengue epidemic, and** $\boldsymbol{t}_{\boldsymbol{e}}$**, the week in which the cumulative cases cross the epidemic threshold value.** $\boldsymbol{\Delta}_{\boldsymbol{t}}$ **is only measured for true positives (that is, in cases where**$\boldsymbol{t}_{\boldsymbol{p}}$ **occurred earlier than** $\boldsymbol{t}_{\boldsymbol{e}}$**).**

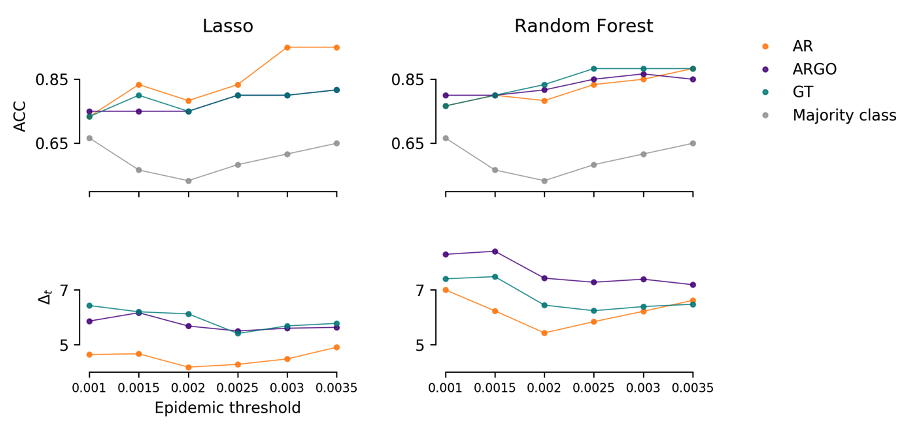

**Figure C. Line plot showing the changes in the label distribution of the time-intervals as a function of the epidemic threshold selected. Common epidemic thresholds such as .001 show cases of imbalanced classes, while the .002 value is closer to a balanced class distribution.**

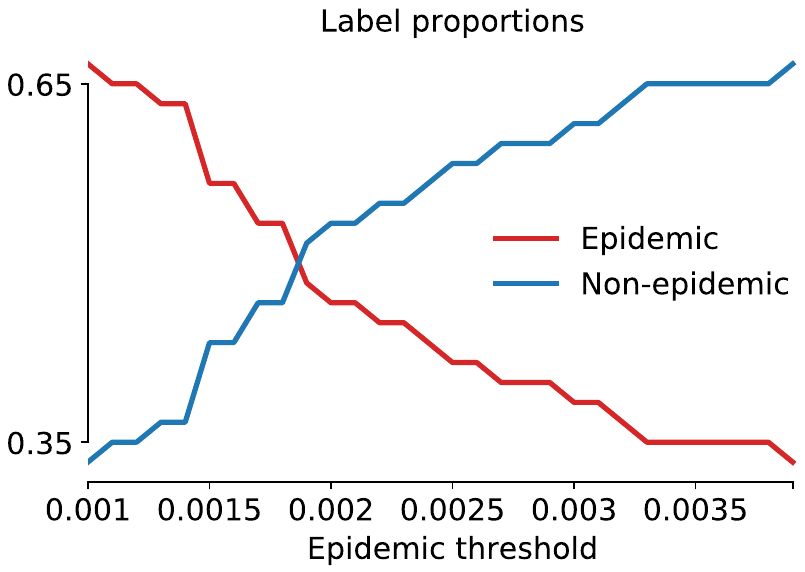

**Plots**

**Figure 1**

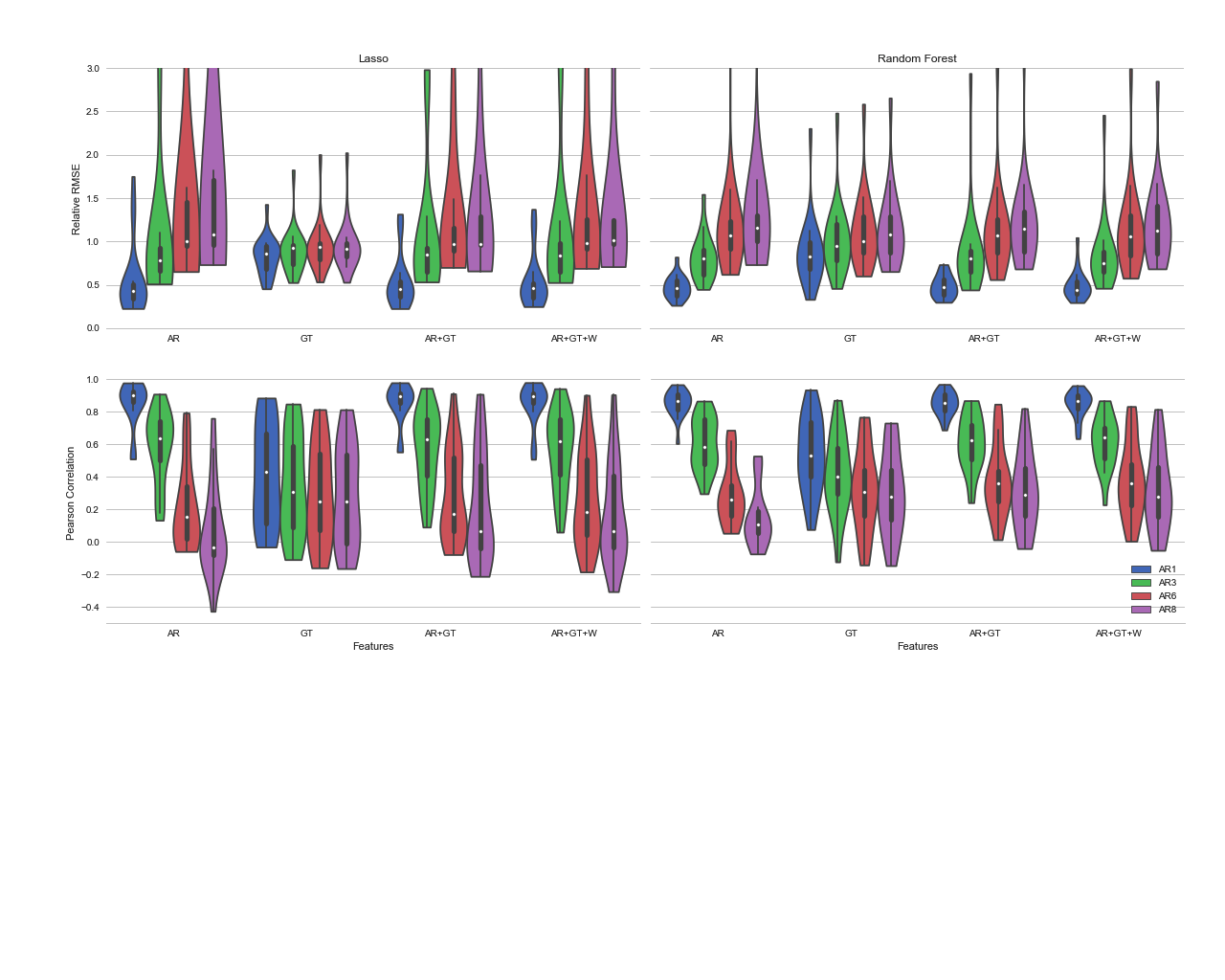

**Figure 2**

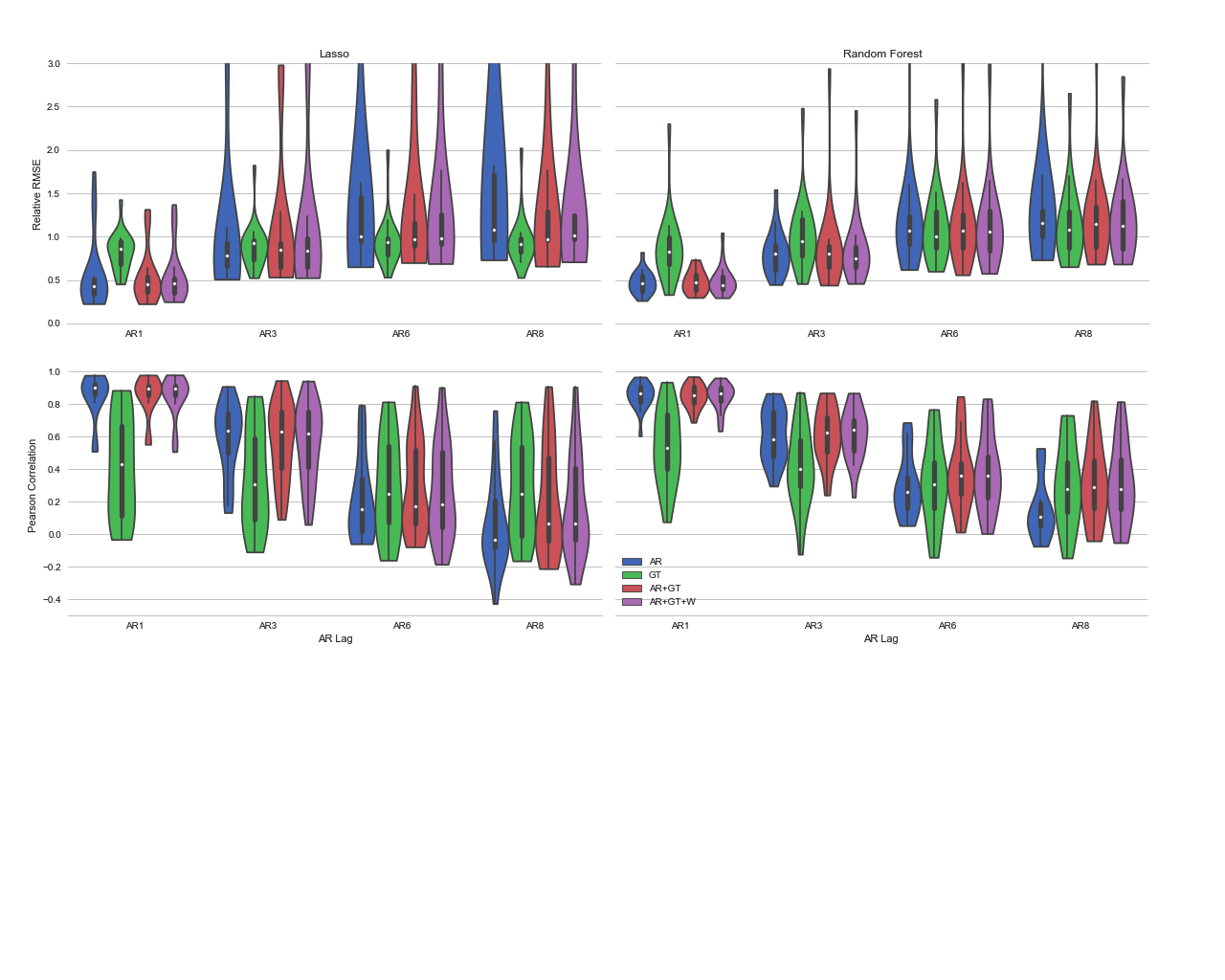

**Figure 3 – Best model for each city, with all time lags**

a. **Feature sets for all lags**

**AR**

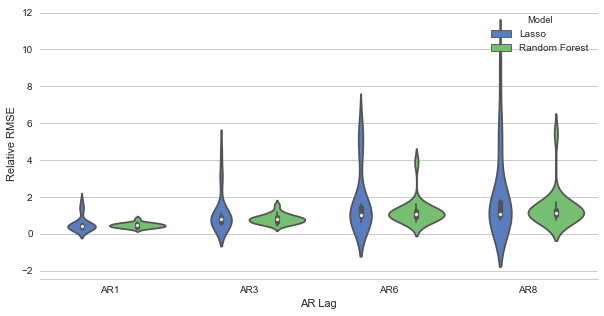

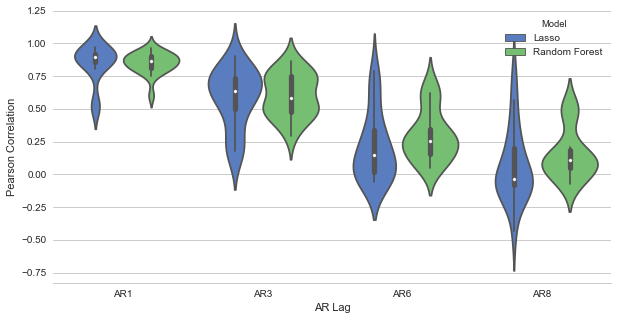

**GT**

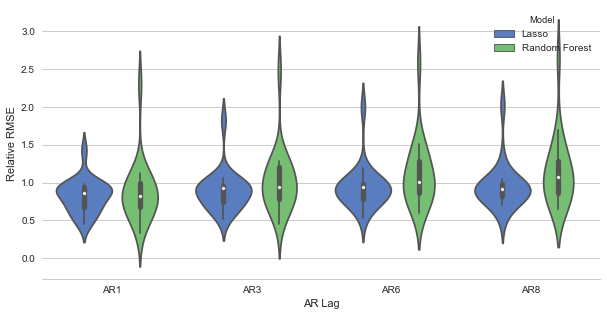

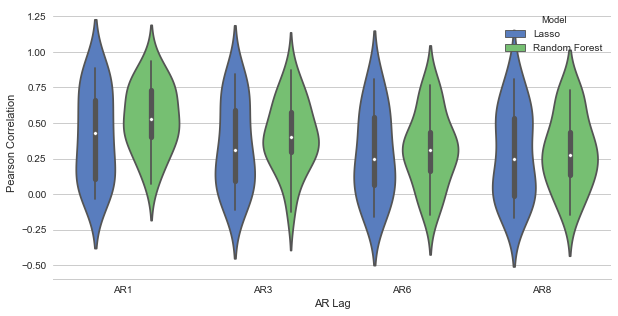

**ARGO**

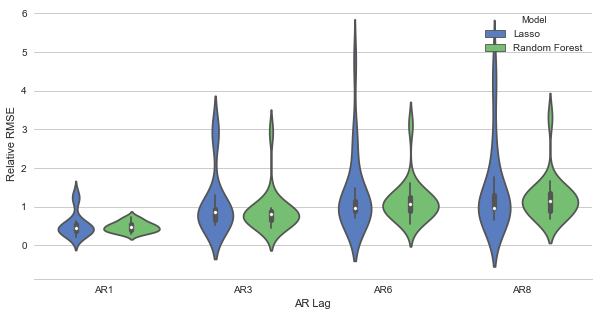

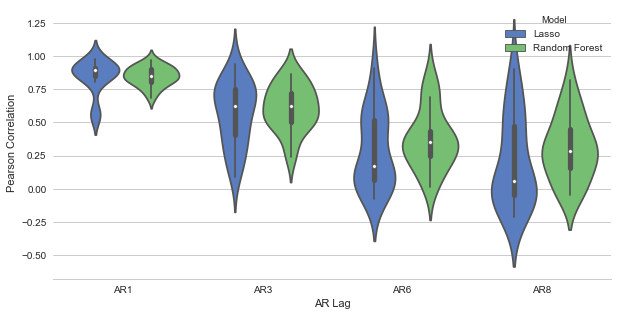

**ARGO + W**

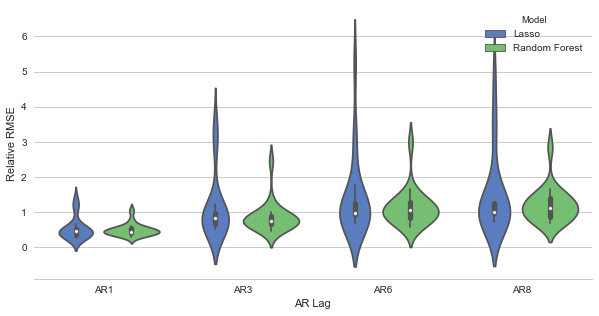

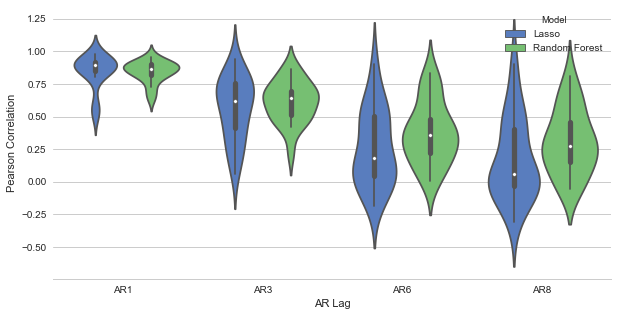

b. **Lags for all feature sets**

**AR1**

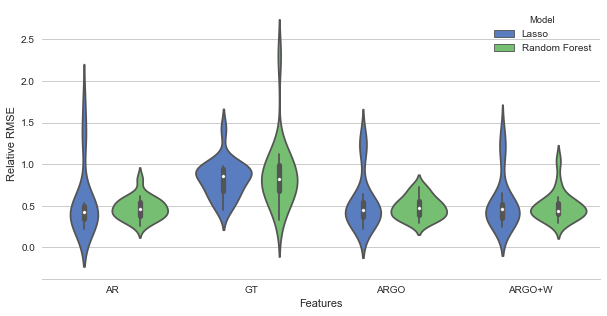

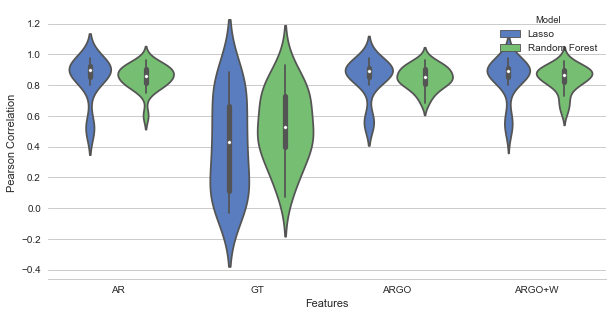

**AR3**

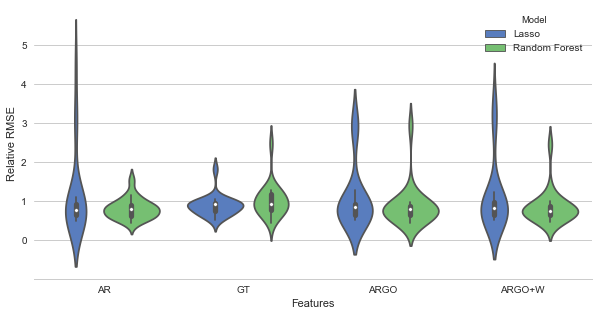

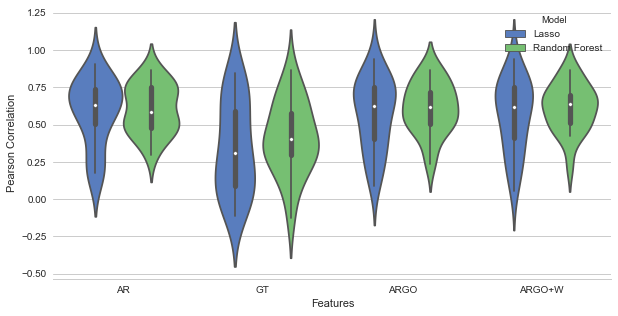

**AR6**

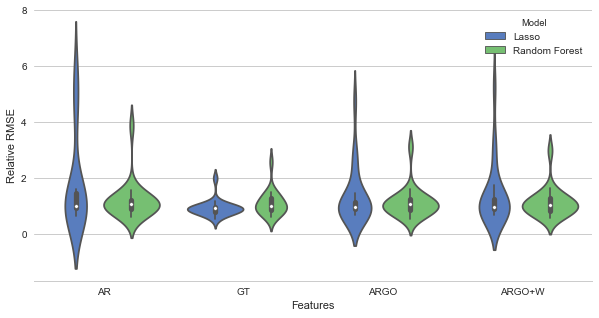

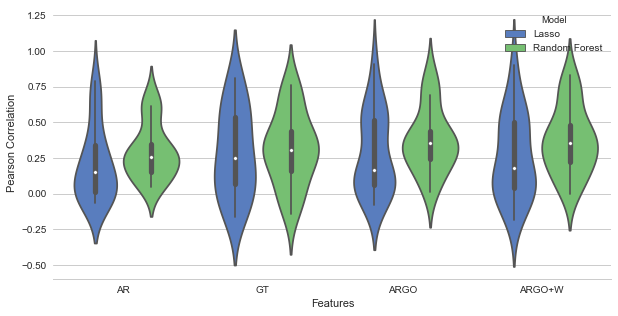

**AR8**

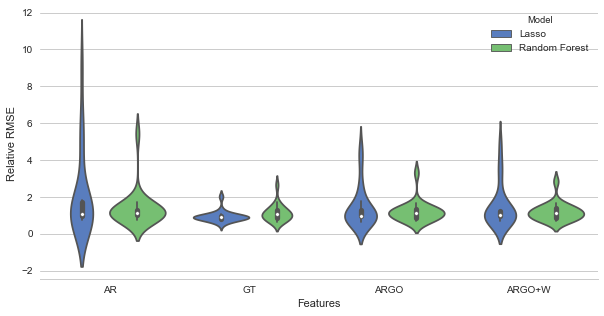

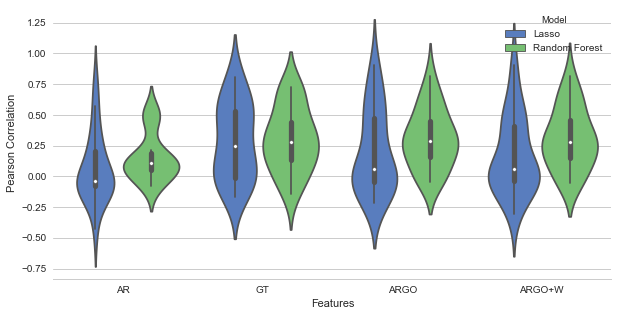

c. **Additional Plots**

**Features with AR Lags as Secondary**

**Lasso**

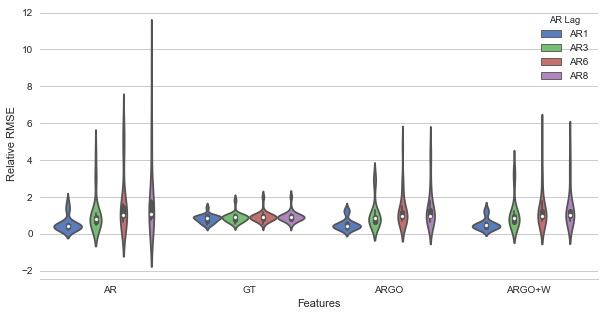

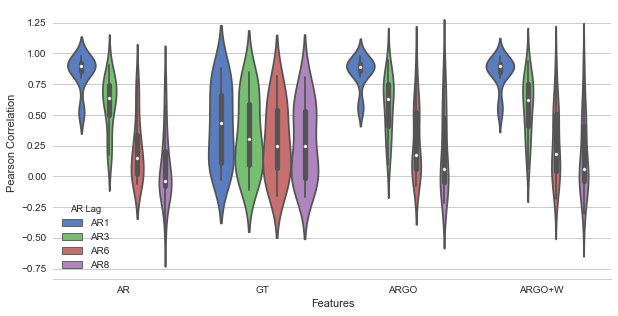

**Random Forest**

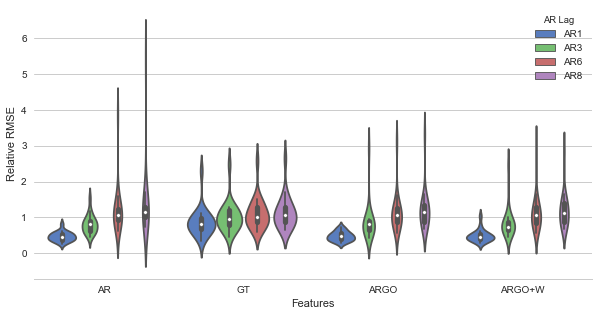

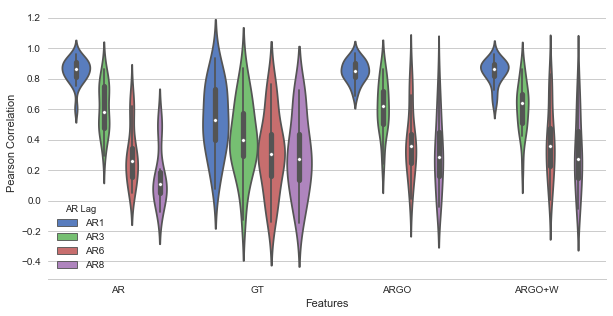

**AR Lags with features as Secondary**

(changed scale here so easier to read)

**Lasso**

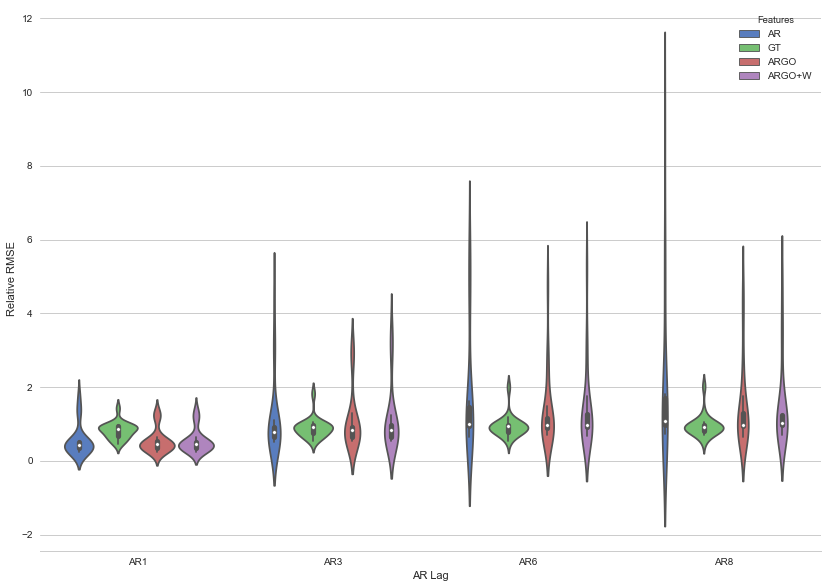

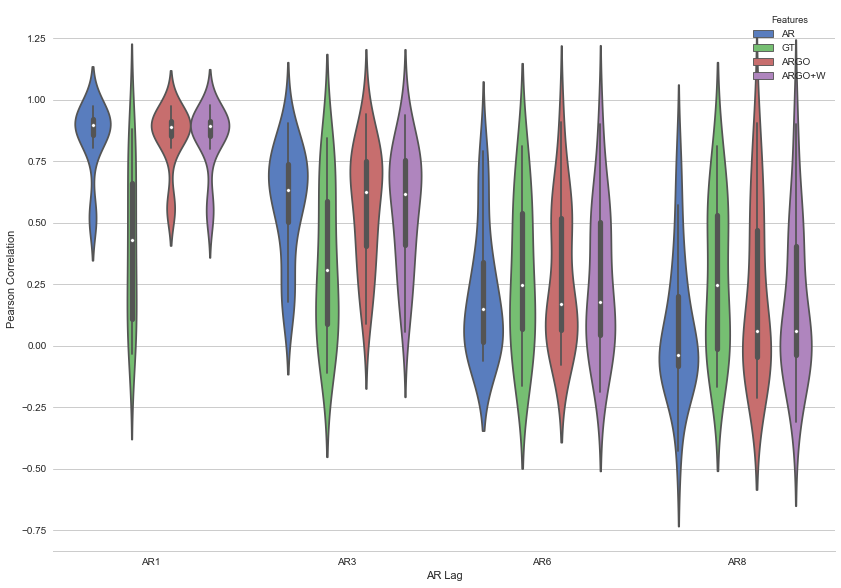

**Random Forest**

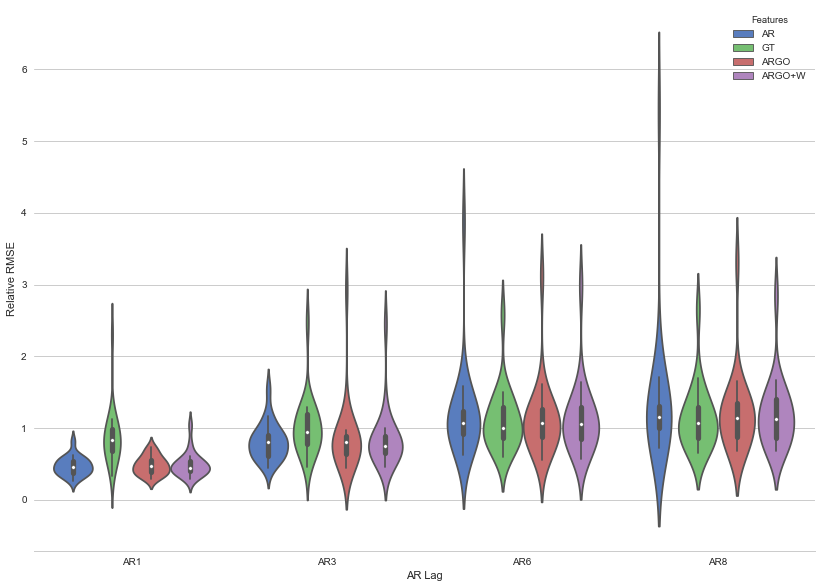

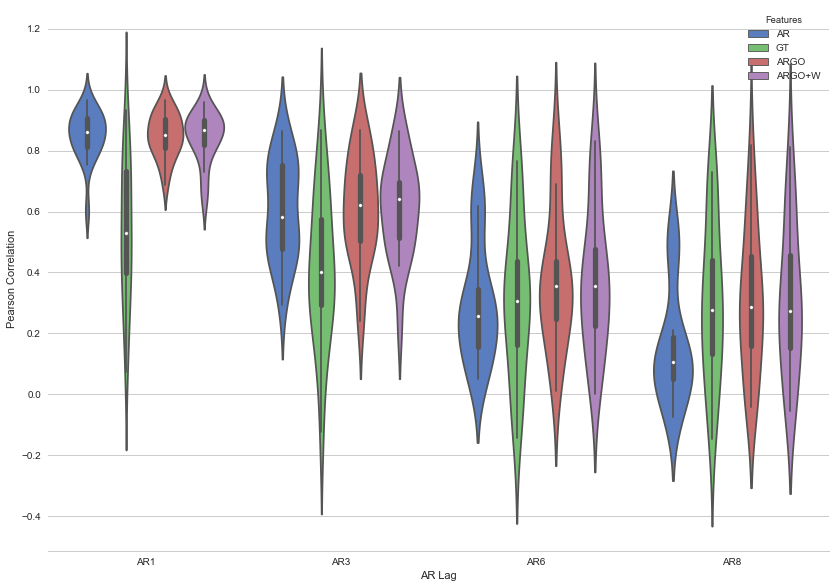

**Appendix: Plots of all cities**

**Comparative Lasso and Random Forest, across all feature sets and AR lags**

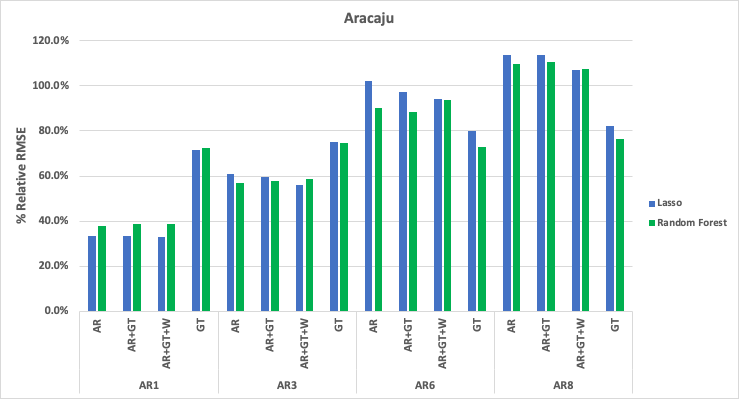

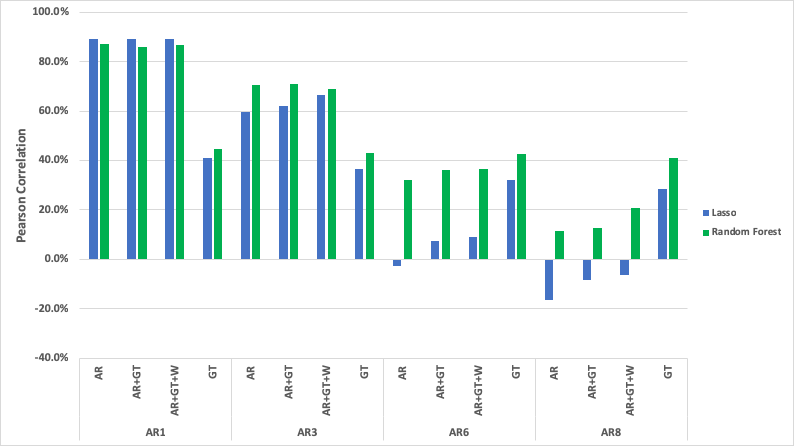

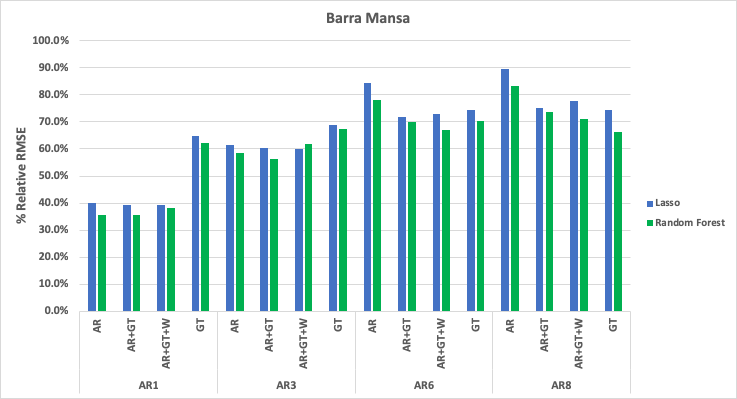

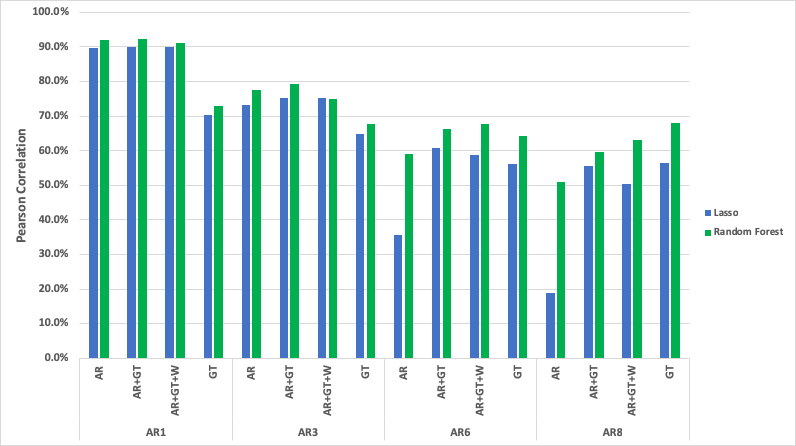

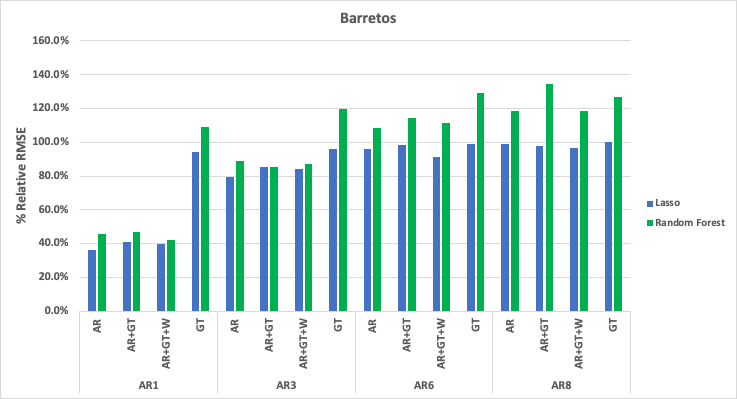

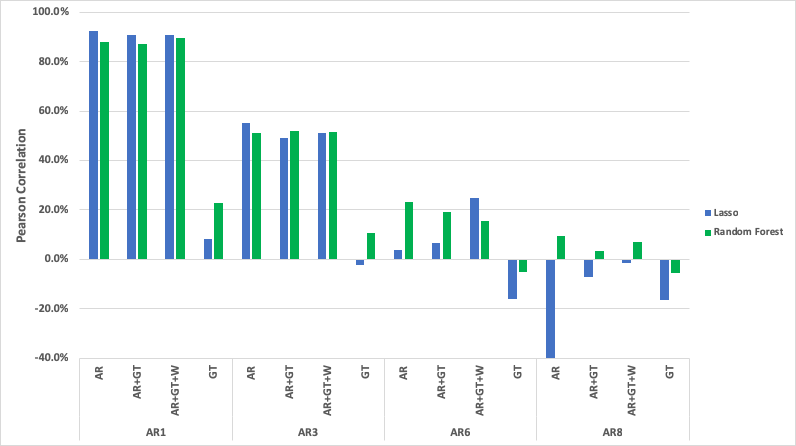

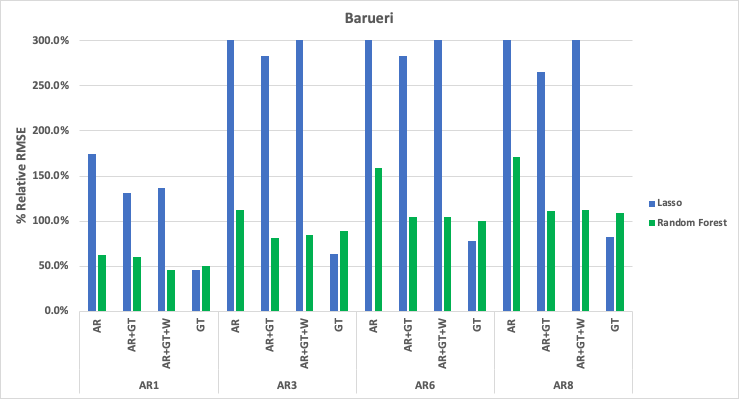

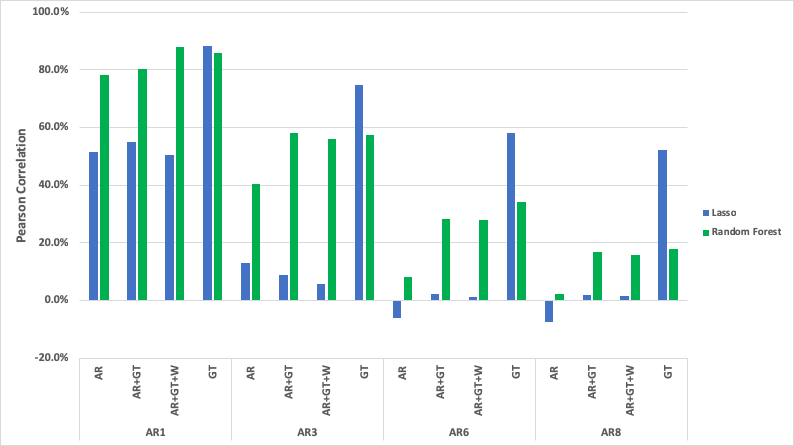

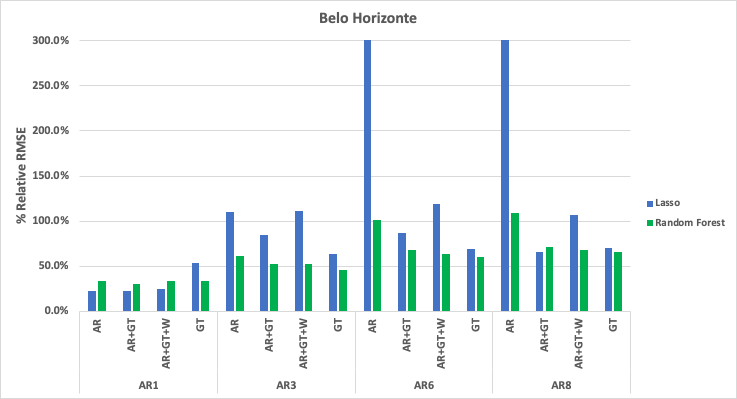

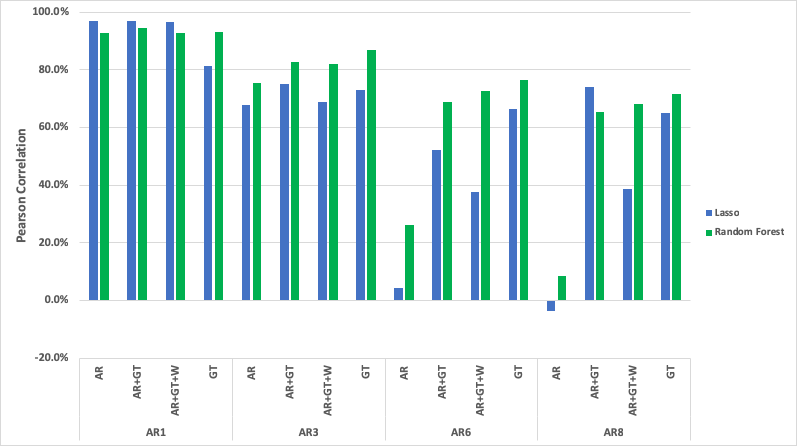

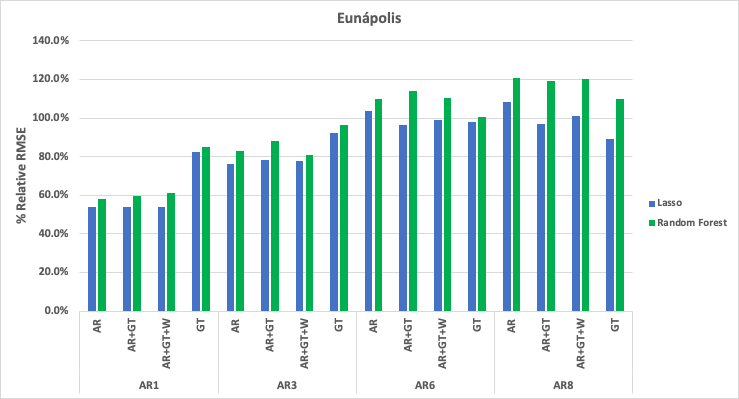

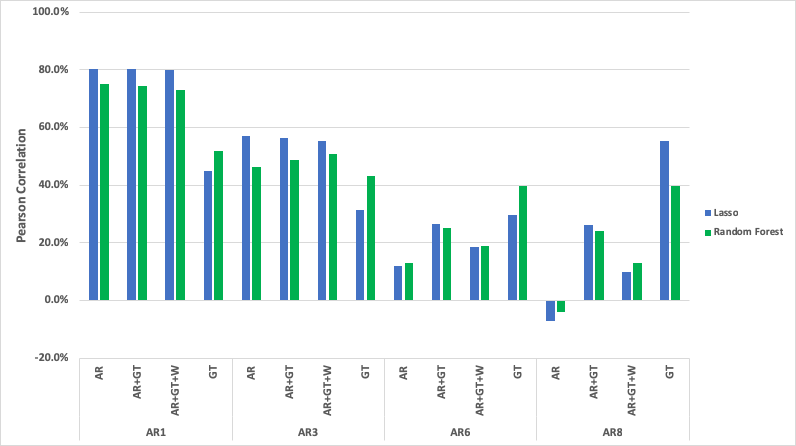

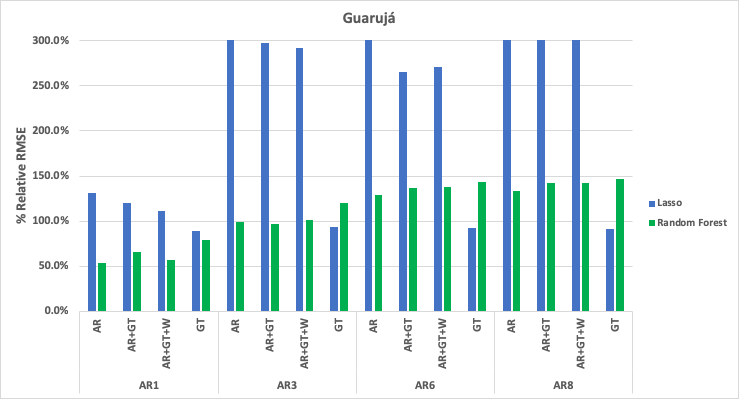

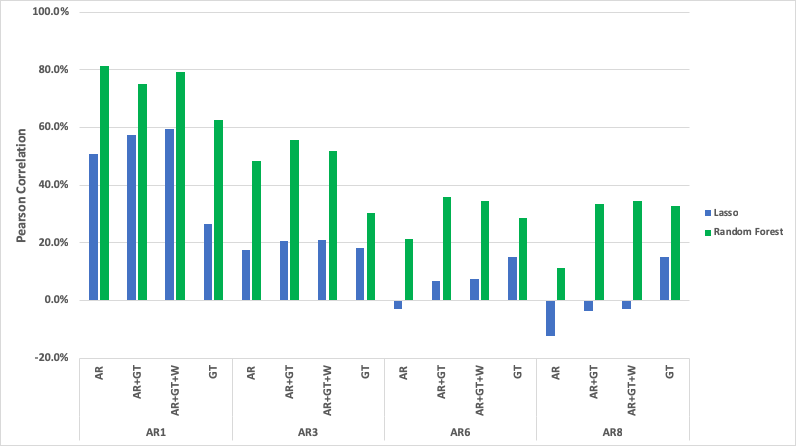

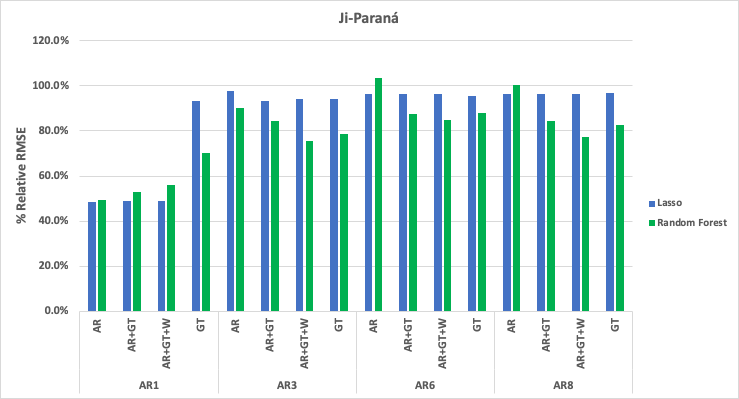

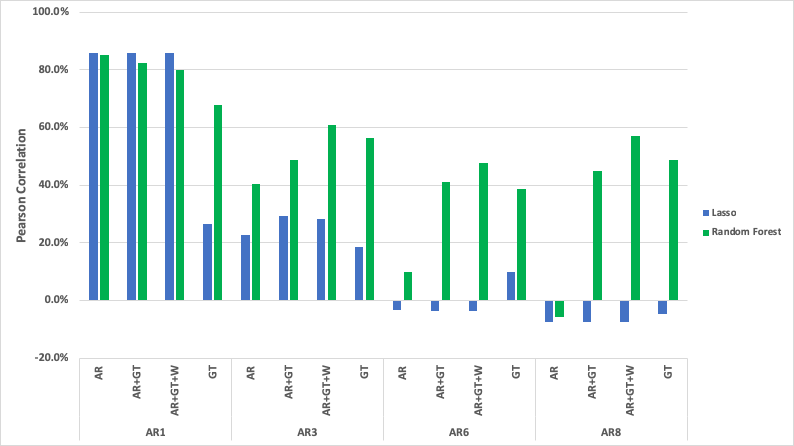

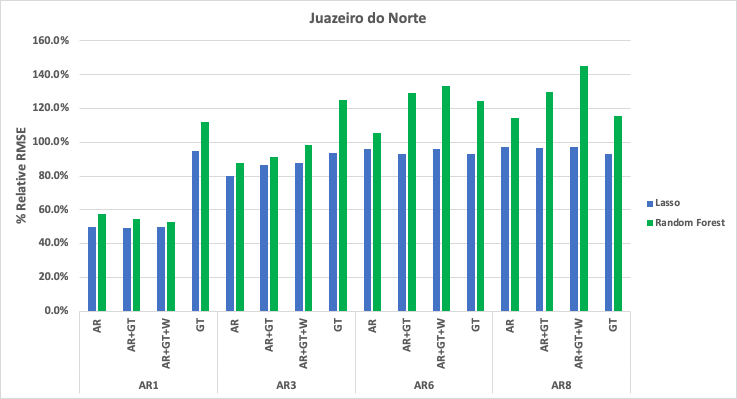

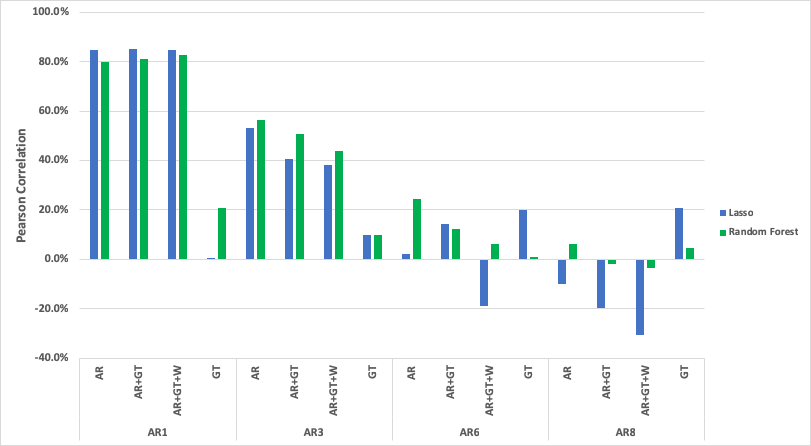

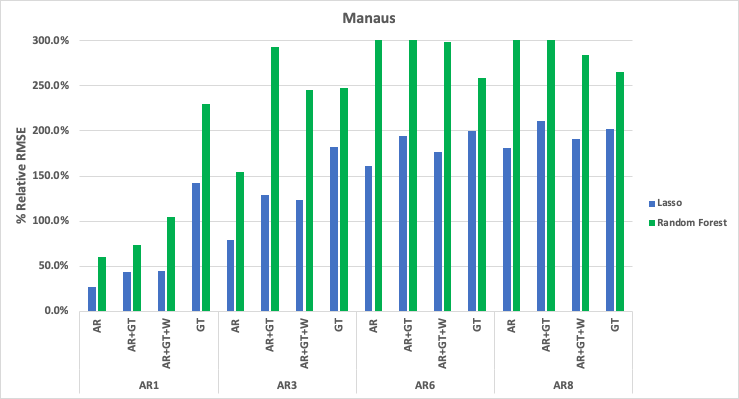

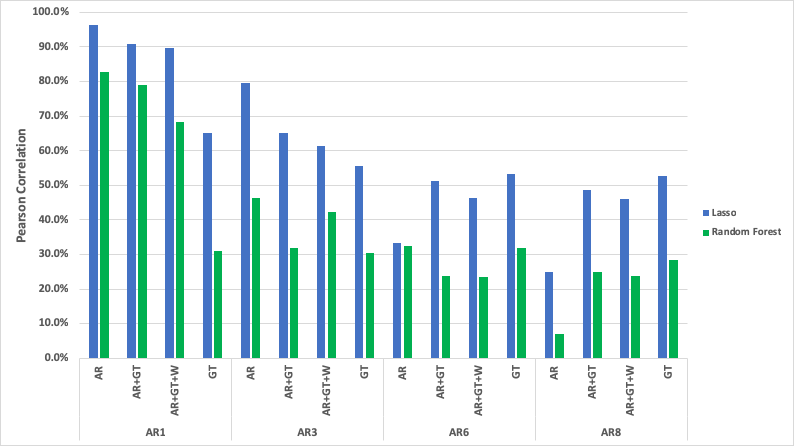

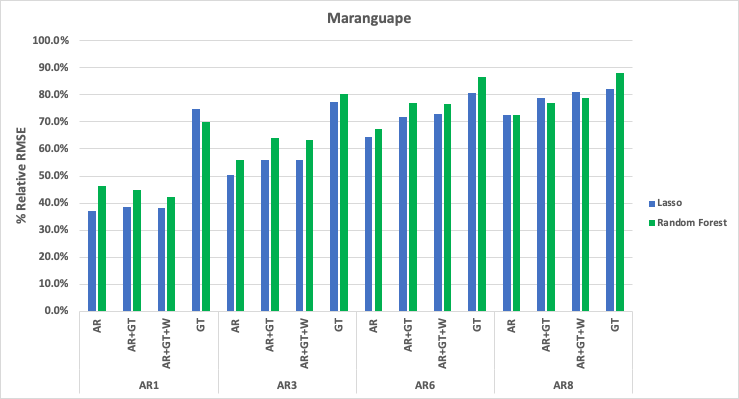

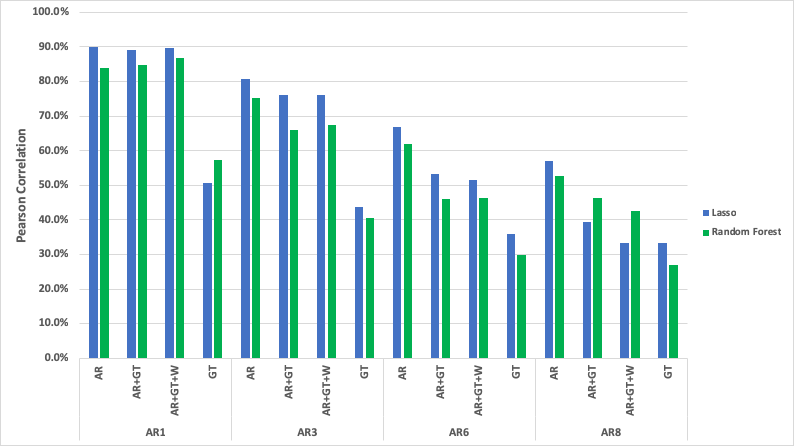

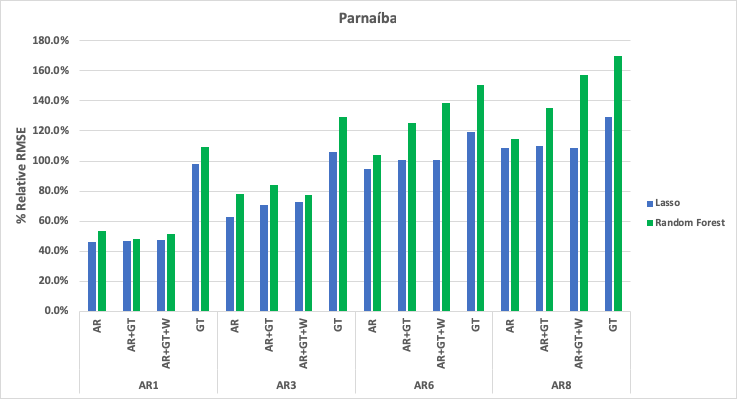

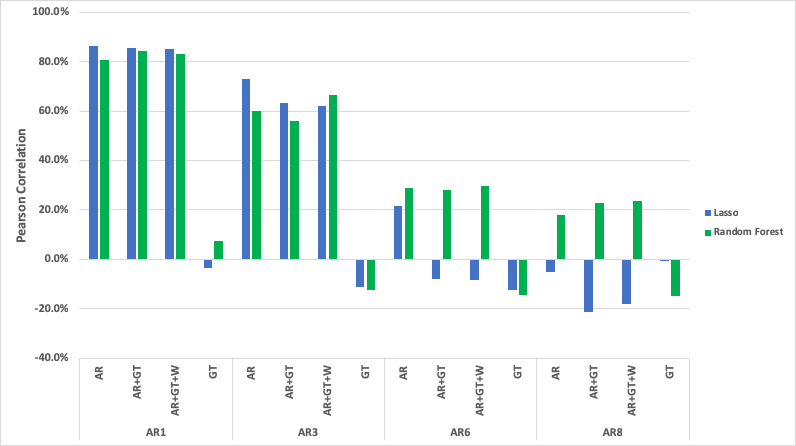

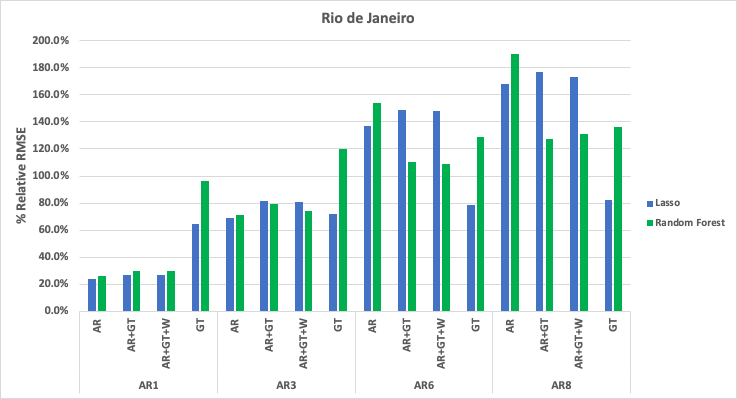

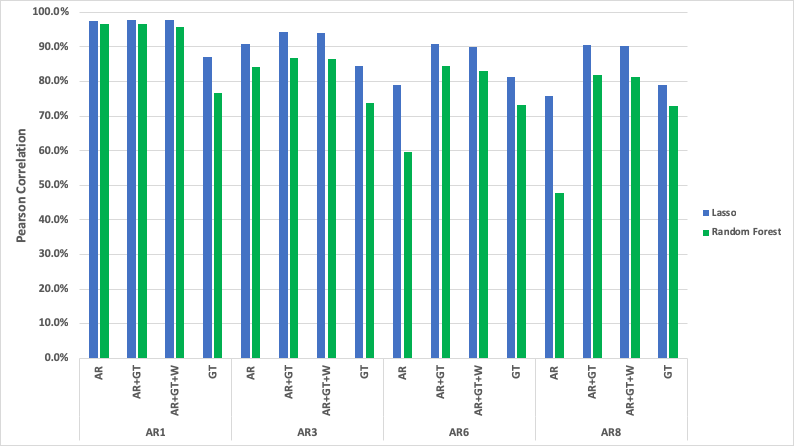

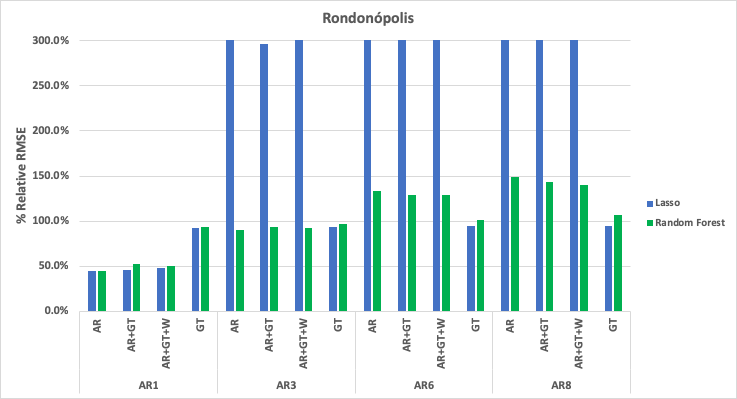

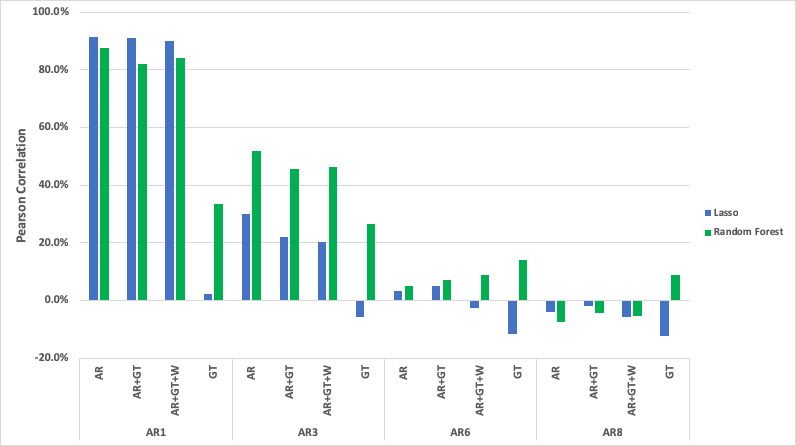

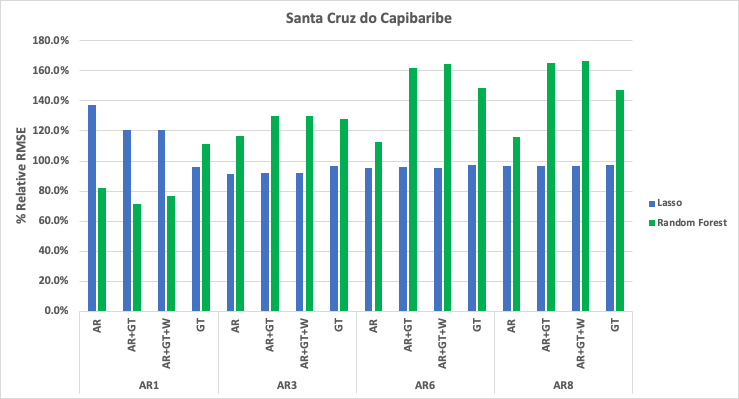

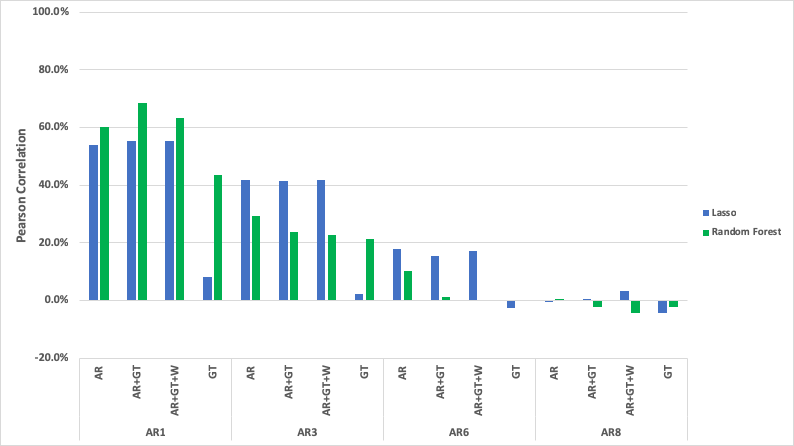

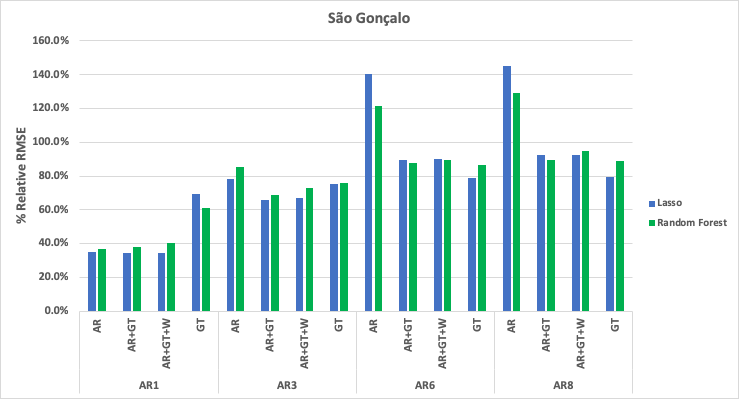

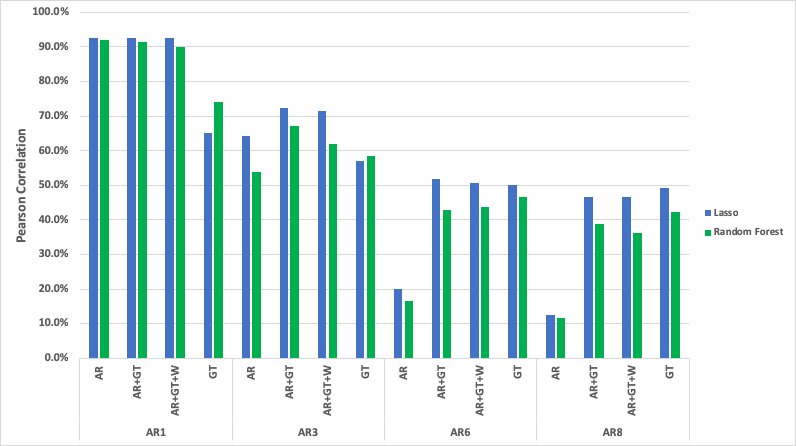

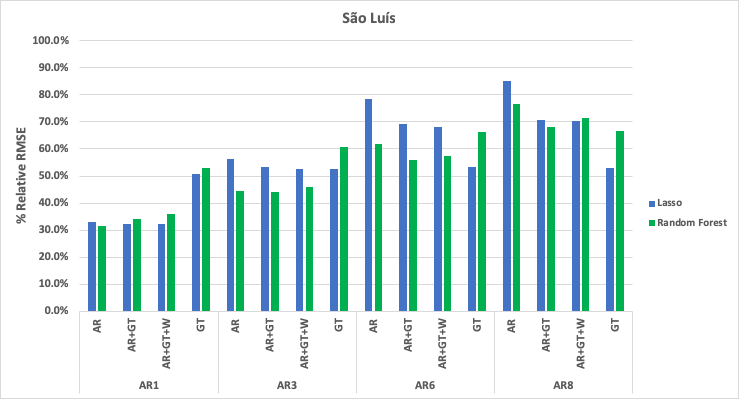

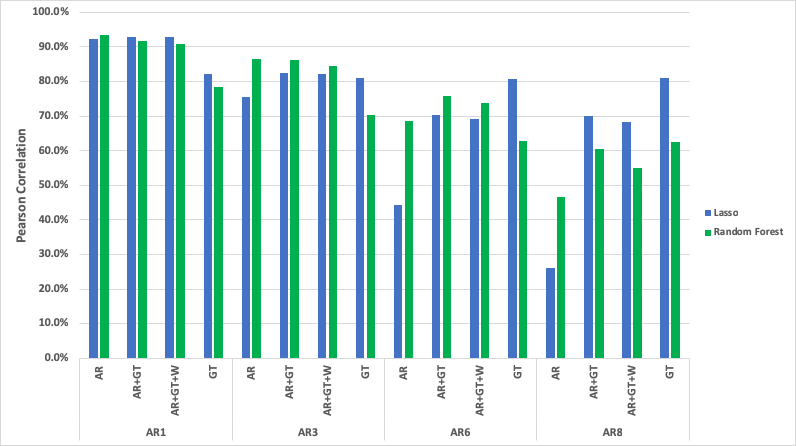

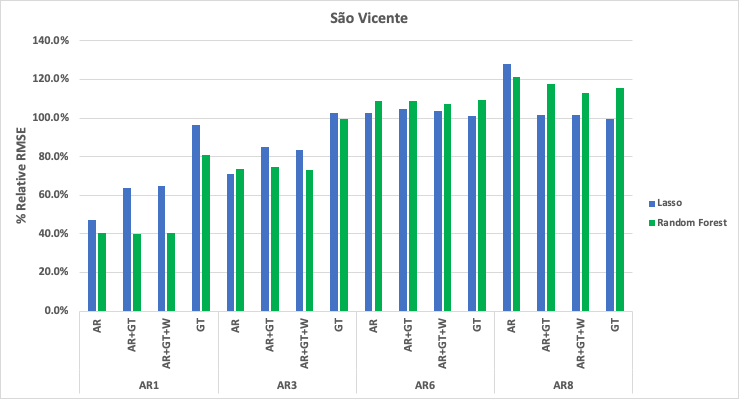

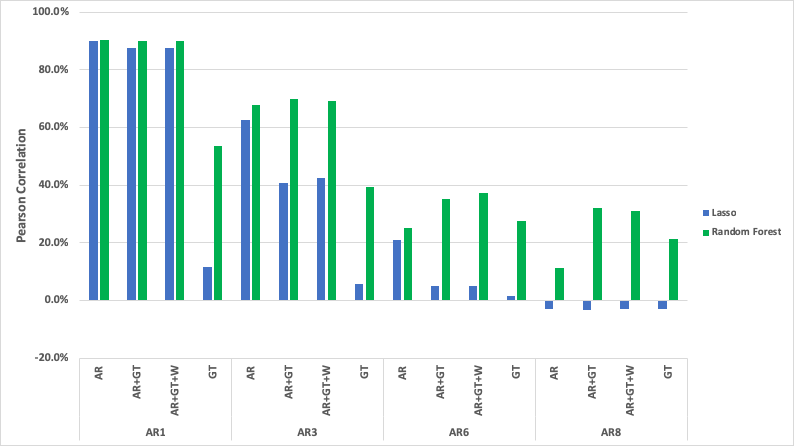

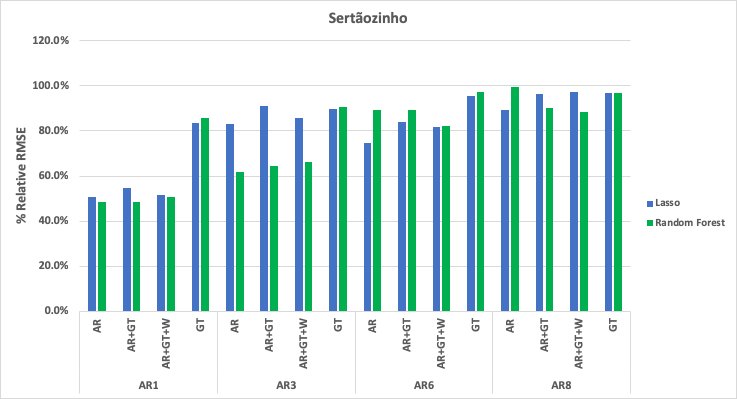

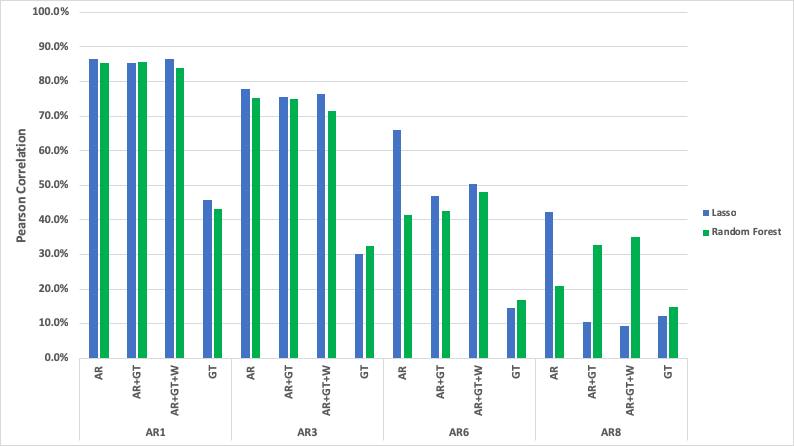

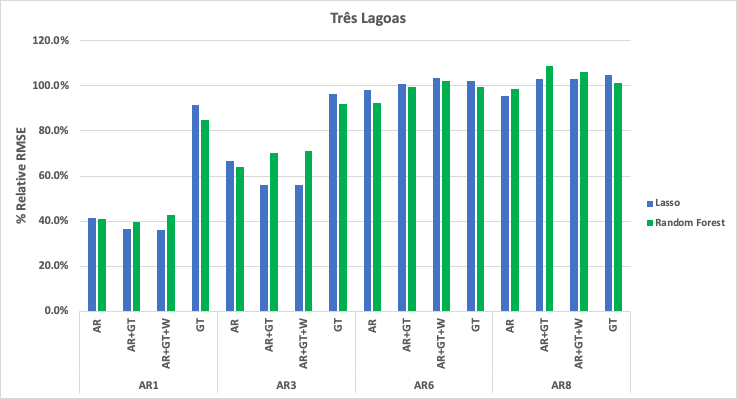

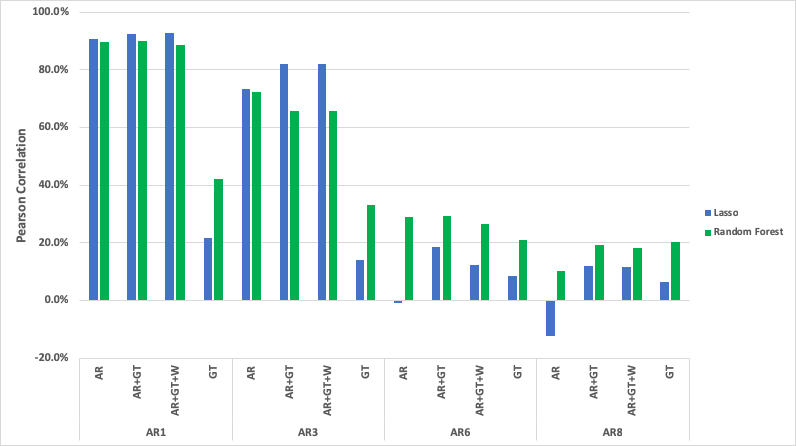
